# A comparison of performance metrics for cloth face masks as source control devices for simulated cough and exhalation aerosols

**DOI:** 10.1101/2021.02.16.21251850

**Authors:** William G. Lindsley, Francoise M. Blachere, Donald H. Beezhold, Brandon F. Law, Raymond C. Derk, Justin M. Hettick, Karen Woodfork, William T. Goldsmith, James R. Harris, Matthew G. Duling, Brenda Boutin, Timothy Nurkiewicz, John D. Noti

## Abstract

Universal mask wearing is recommended by the Centers for Disease Control and Prevention to help control the spread of COVID-19. Masks reduce the expulsion of respiratory aerosols (called source control) and offer some protection to the wearer. However, masks vary greatly in their designs and construction materials, and it is not clear which are most effective. Our study tested 15 reusable cloth masks (which included face masks, neck gaiters, and bandanas), two medical masks, and two N95 filtering facepiece respirators as source control devices for aerosols ≤ 7 µm produced during simulated coughing and exhalation. These measurements were compared with the mask filtration efficiencies, airflow resistances, and fit factors. The source control collection efficiencies for the cloth masks ranged from 17% to 71% for coughing and 35% to 66% for exhalation. The filtration efficiencies of the cloth masks ranged from 1.4% to 98%, while the fit factors were 1.3 to 7.4 on an elastomeric manikin headform and 1.0 to 4.0 on human test subjects. The correlation coefficients between the source control efficacies and the other performance metrics ranged from 0.31 to 0.66 and were significant in all but one case. However, none of the alternative metrics were strong predictors of the source control performance of cloth masks. Our results suggest that a better understanding of the relationships between source control performance and metrics like filtration efficiency, airflow resistance, and fit factor are needed to develop simple methods to estimate the effectiveness of masks as source control devices for respiratory aerosols.

## Introduction

Humans infected with SARS-CoV-2, the virus that causes coronavirus disease 2019 (COVID-19), can produce droplets and aerosols of respiratory fluids containing the virus when they cough, breathe, talk, sing and sneeze (Anderson et al. 2020; CDC 2020d; Hamner et al. 2020; Ma et al. 2020; Morawska and Milton 2020). To reduce the transmission of SARS-CoV-2, public health agencies have recommended that the general public wear cloth masks (CDC 2020a; b; 2021; Edelstein and Ramakrishnan 2020; WHO 2020). The primary purpose of masks, which includes face masks, neck gaiters, bandanas and other face coverings, is to block the expulsion of infectious droplets and aerosols from the wearer into the environment (called source control) and thereby reduce the exposure of other people to the virus (CDC 2020c). Laboratory studies using manikins and human subjects have shown that cloth face masks can partially block respiratory aerosols produced during coughing, breathing and talking (Asadi et al. 2020; Davies et al. 2013; Lindsley et al. 2021; Pan et al. 2020). Wearing medical face masks (i.e., ‘surgical masks’ as defined by the U.S. Food and Drug Administration (FDA 2004)) reduces the dispersion of potentially infectious aerosols from patients with respiratory infections (Leung et al. 2020; Milton et al. 2013). Masks may also provide some personal protection to the wearer by reducing their exposure to infectious droplets and aerosols produced by others, although they are not as effective as a respiratory protective device such as a NIOSH-approved N95 filtering facepiece respirator (N95 respirator) (CDC 2020c; Lawrence et al. 2006; Oberg and Brosseau 2008; Pan et al. 2020). Several community level studies have shown that universal masking helps reduce the spread of COVID-19 (CDC 2020c). For example, a comparison of counties in the US state of Kansas found that mask mandates were associated with lower incidence rates of COVID-19 (Van Dyke et al. 2020). A study of 10 US states found that statewide mask mandates were associated with a decline in weekly COVID-19–associated hospitalization growth compared to states without such mandates (Joo et al. 2021).

In response to the need for source control devices for the general public, manufacturers worldwide have produced a broad array of masks. Unfortunately, because of the many different designs and construction materials, it is not possible to predict how well a particular mask will perform as a source control device without testing, which is rarely done for non-medical masks not intended for occupational use. Although general guidelines have been developed (CDC 2020b), it is very difficult for public health organizations, governments, medical facilities, and the general public to know which of the available devices are most effective.

Test methods and performance standards do exist for regulated medical face masks and respiratory protective devices (Rengasamy et al. 2017). In the United States, respiratory protective devices, which are devices such as N95 filtering facepiece respirators that are intended to protect the wearer from airborne particles, must be approved by the National Institute for Occupational Safety and Health (NIOSH) under 42 Code of Federal Regulations (CFR) Part 84 (NIOSH 1995). The approval process includes testing the filtration efficiency for the most-penetrating aerosol particle size and measuring the airflow resistance of the device (NIOSH 2019). Medical face masks that are not intended to be used as respiratory protective devices are cleared by the Food and Drug Administration (FDA), which reviews manufacturer-supplied information on filtration efficiency, airflow resistance, resistance to fluid penetration and flammability (FDA 2004). The FDA recommends that manufacturers use the ASTM standards F2299 for measuring particle filtration efficiency through the mask material using latex microspheres (ASTM 2003), and the F2101 Standard test method of evaluating the bacterial filtration efficiency (BFE) of medical face mask materials, using a biological aerosol of *Staphylococcus aureus* (ASTM 2019). Bacterial filtration efficiency can also be measured with the modified Greene and Vesley method using human test subjects (Greene and Vesley 1962; Quesnel 1975).

In addition to filtration performance, how well a respiratory protective device protects the wearer depends upon how well the device fits the face and whether the seal between the face and the device has gaps or leaks (Lawrence et al. 2006). ASTM Standard F3407-20 outlines procedures for testing respirators using a bivariate panel of human test subjects with different facial dimensions (ASTM 2020). The NIOSH approval test measures the filtration properties of respirators and masks but does not include tests of how well the device fits faces of different shapes, although this has been proposed. Consequently, in the United States, when a worker is required to wear a respirator, the Occupational Safety and Health Administration (OSHA) requires that a respirator fit test be performed annually for each worker to determine how well the respirator protects the worker (OSHA 2020). Quantitative fit tests measure the aerosol particle concentration inside and outside the respirator during a series of exercises, and this information is used to calculate the fit factor (outside concentration/inside concentration) (Janssen and McKay 2017). The minimum acceptable fit factor for a respirator depends upon the exposure level and potential health consequences of the hazards to which the worker may be exposed (OSHA 2020).

The existing test methods for respirators and medical face masks provide a possible basis for developing standards for non-medical masks used as source control devices. The American Association of Textile Chemists and Colorists (AATCC) published guidelines for cloth face coverings that include the use of ASTM F2299 and other national and international performance standards (AATCC 2020). ASTM International is also developing a specification for face coverings (ASTM 2021). To assist in the development of these types of standards and guidelines, the relationship between the efficacy of masks as source control devices as compared to their efficacy using different performance measures needs to be better understood. For example, it is possible to measure the filtration efficiency of the mask material, but it is not clear how that filtration efficiency translates to source control efficacy or what minimum performance level should be required.

The purpose of this study was to compare the source control performance of a variety of N95 respirators, medical masks, and cloth masks with the filtration efficiency, airflow resistance, fit factor measured using manikin headforms, and fit factor measured on humans. The results of these experiments will assist in the development of appropriate test methods and standardized performance metrics to evaluate the efficacy of cloth masks as source control devices for respiratory aerosols.

## Materials & methods

### Terminology

In our study, “cloth masks” refers to cloth face masks, neck gaiters and bandanas. The term “cloth mask” applies to any mask constructed from textiles or fabrics (both natural and synthetic) that is not a surgical mask or N95 respirator and is not intended for use as personal protective equipment. The term “surgical mask” applies to commercially produced masks regulated by the U.S. Food and Drug Administration under 21 CFR 878.4040 for performing medical procedures (FDA 2004). A surgical mask covers the user’s nose and mouth and provides a physical barrier to fluids and particulate materials. “Surgical masks” may include masks that are labeled as a surgical, laser, isolation, dental or medical procedure masks with or without a face shield. Surgical masks may be variably shaped, (e.g., duck bill, flat pleated, cone shaped, pouch). They are loose-fit and are not expected to provide as reliable a level of protection against airborne or aerosolized particles as N95 respirators regulated by NIOSH.

### Experimental design

A source control measurement system was used to assess the efficacy of N95 respirators, medical masks, and cloth masks as source control devices for respiratory aerosols. The source control performance was determined by measuring the collection efficiency of the mask, which is the fraction of the mass of the coughed or exhaled test aerosol particles that were blocked by the mask from reaching the collection chamber. Fit tests were performed both on a manikin headform with pliable skin and with human test subjects. The filtration efficiencies and airflow resistance of the construction materials were measured using a modified version of the test method used for respirator approvals. The source control experiment results were then compared with the fit factors, filtration efficiencies and airflow resistances measured on the same devices.

### Respiratory aerosol source control measurement system

The efficacy of respirators, medical masks, and cloth masks as source control devices for aerosols produced during coughing and exhalation was determined using a respiratory aerosol source control measurement system described previously (Lindsley et al. 2021). The system includes a coughing and breathing aerosol simulator, a manikin headform, an aerosol collection chamber, and a cascade impactor (Figure 1). The manikin headform used in the study has pliable skin that mimics the elastic properties of human skin in order to create a realistic simulation of how each source control device would fit a human face (Bergman et al. 2014).

**Figure 1:**
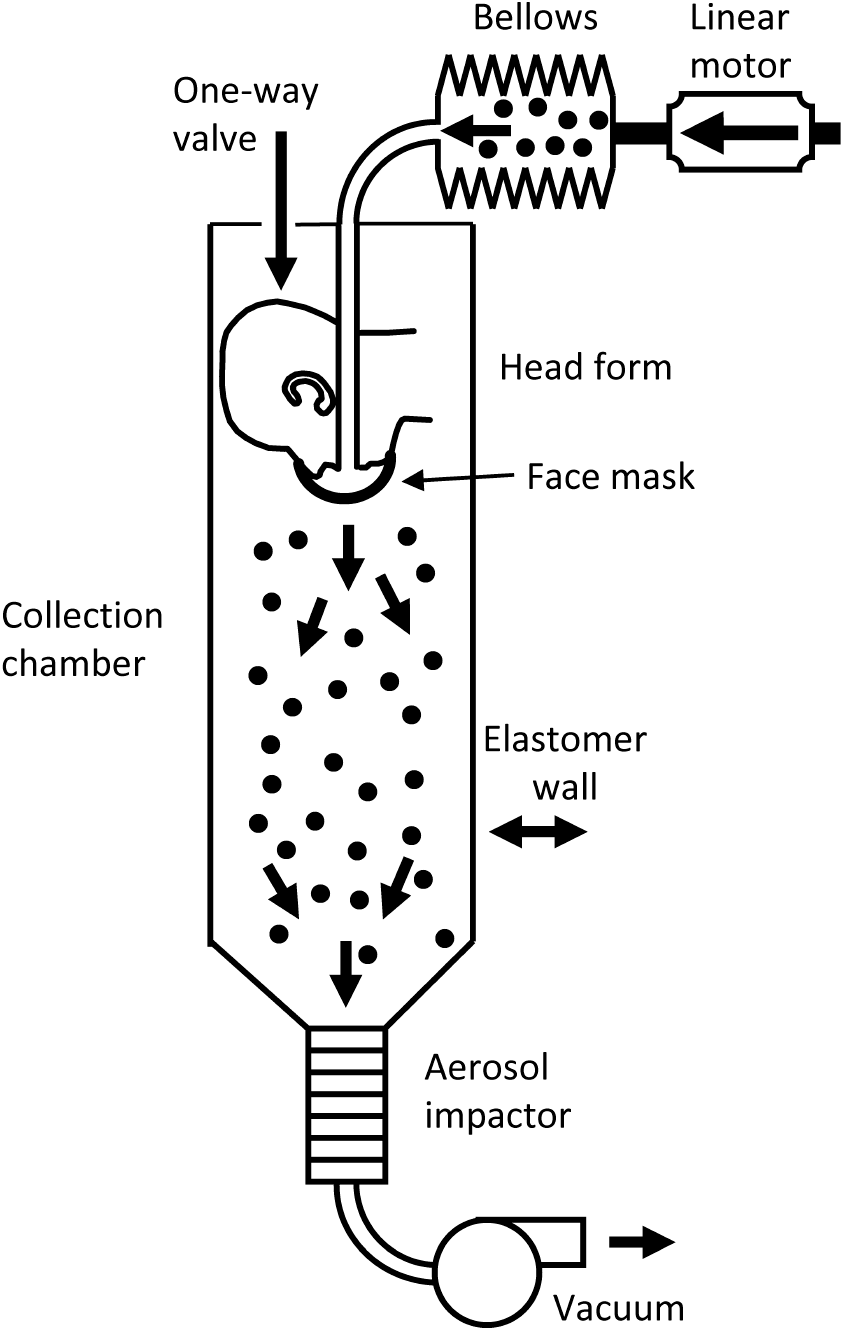
Source control measurement system. The system consists of an aerosol generation system, a bellows and linear motor to produce the simulated coughing and breathing, a pliable skin headform on which the source control device is placed, a 105 liter collection chamber into which the aerosol is coughed or exhaled, and a cascade impactor to separate the aerosol particles by size and collect them. The system is described in more detail in Lindsley et al. (2021).

The test aerosol was produced using a solution of 14% potassium chloride (KCl) and 0.4% sodium fluorescein in a single-jet Collison nebulizer (BGI, Butler, NJ) at 103 kPa (15 lbs./in^2^). The aerosol passed through a diffusion drier (Model 3062, TSI, Shoreview, MN), mixed with dry filtered air flowing at 10 L/min for the cough tests and 15 L/min for the breathing tests, and neutralized using a bipolar ionizer (Model HPX-1, Electrostatics). An elastomeric bellows driven by a computer-controlled linear motor produced the coughing or breathing airflow.

For cough tests, the test aerosol was loaded into the elastomeric bellows and then coughed out using a single cough with a volume of 4.2 L and a peak flow rate of 11 L/s (Lindsley et al. 2013). An Andersen cascade impactor (Model TE-10-830, Tisch Environmental) collected all aerosol particles that traveled through or around the device for 20 minutes after each cough. The impactor operated at a flow rate of 28.3 liters/minute and had six collection stages and a filter that separated the aerosol particles into seven size fractions based on the aerodynamic diameter of the particles: <0.6 µm; 0.6-1.1 µm; 1.1-2.1 µm; 2.1-3.3 µm; 3.3-4.7 µm; 4.7-7.0 µm; and >7 µm. The impactor collection plates were coated with a solution of glycerol and Brij 35 to prevent particles from bouncing off the plates during collection (Mitchell 2003). Because the amount of aerosol in the largest size fraction was small (<0.7% of total aerosol mass) and because of possible losses due to settling of the large aerosol particles, data for the largest size fraction was not included in the analysis.

For breathing tests, the system used a ventilation rate of 15 L/min with a breathing rate of 12 breathes/min and a tidal volume of 1.25 liters, which corresponds to the ISO standard for a female performing light work (ISO 2015). The test aerosol was only generated for the first 30 seconds of breathing to avoid overloading the impactor. The breathing continued for 20 minutes total. The impactor collected the aerosol particles in the chamber during the 20-minute breathing period followed by an additional five minutes of collection after the breathing had stopped.

### Respirators, Medical Masks, and Cloth Masks

Nineteen commercially-available N95 respirators, medical masks, and cloth masks were selected to provide a broad cross-section of the different types of source control devices that are available (Table 1). For the source control tests, each device was placed on the headform as it would normally be worn by a person. Three of the neck gaiters were tested both as a single layer of fabric and folded over to provide two layers of fabric. Before the source control test, the manikin fit factor (Janssen and McKay 2017) was measured by performing a respirator fit test (Bergman et al. 2015) for each device using a PortaCount® Pro+ respirator fit tester (Model 8038, TSI, Shoreview, MN) in N95 mode with the system breathing at 36 L/min but not producing an aerosol. Each device was used for two consecutive tests. Photographs of the source control devices on the headform are shown in the online supplementary information (SI; Figures S1-S3).

**Table 1:** Source control devices used in experiments. In this paper, the reusable cloth face masks, neck gaiters and bandanas are referred to collectively as “cloth masks”. IL is inner layer, ML is middle layer, and OL is outer layer. Photographs are shown in Figures S1-S3.

| *Type* | *Manufacturer* | *Product* | *Fastener* | *Material* |
| --- | --- | --- | --- | --- |
| Medical masks & respirators | 3M | Model 1860 N95 respirator and surgical mask (TC-84A-0006) | Two elastic straps around head | Polypropylene and polyethylene composition |
|  | BYD | N95 filtering facepiece respirator (TC-84A-9221) | Two elastic straps around head | IL and OL: Polypropylene spun-bound nonwoven fabric. ML: Polypropylene melt-blown nonwoven + hot air cotton |
|  | 3M | Model 1818 surgical mask | Two ties behind head | Poly(ethylene terephthalate) and polypropylene |
|  | Excellent Artisan | Procedure mask | Ear loops | 3-ply, Nonwoven. Fabric composition unknown |
| Reusable cloth face masks | Hanes | Defender 3-ply mask | Ear loops | 100% cotton |
|  | EnerPlex | 3-ply mask | Ear loops | IL and ML: 65% polyester, 35% cotton. OL: 100% polyester |
|  | Craft & Soul | 2-ply mask | Ear loops | 65% polyester, 35% cotton |
|  | Lefty | 2-ply face mask | Ear loops | 92% polyester, 8% spandex |
|  | Badger | 4-ply mask | Ear loops | IL : 100% cotton. MLs: melt blown filters. OL: air mesh polyester |
|  | Debrief Me | 3-ply mask with PM 2.5 filter | Ear loops | IL 100% cotton, ML and OL: polyester blend |
|  | Fabrique Innovations | Mask | Ear loops | 100% cotton |
|  | Besungo | Mask with PM 2.5 filter & exhalation valves | Ear loops and Velcro neck strap | Nylon |
|  | Inspire | 2-ply mask with PM 2.5 filter | Ear loops | Polyester/cotton blend |
| Neck gaiters | FKGIONG | Neck gaiter | Encircles head | 95% polyester, 5% spandex |
|  | AXBXCX | Neck gaiter | Encircles head | Polyester |
|  | Retro Brand | Neck gaiter | Encircles head | 100% polyester |
|  | Givon | Neck gaiter | Encircles head | 100% polyester fleece |
| Bandanas | L & M | Bandana | Ties behind head | 100% cotton |
|  | Underwear | Bandana/gaiter | Ear loops | Polyester blend |

### Filtration efficiency and airflow resistance measurements

The filtration efficiency and airflow resistance of the construction materials were measured using automated filter testers (Models 8130 and 8130A, TSI). Material samples were secured to a test plate using beeswax as shown in Figure S4 in the SI. Measurements were made using a modified version of the NIOSH standard testing procedure (STP) (NIOSH 2019). Under the modified STP, samples were tested at ambient temperature and humidity but were not subjected to conditioning at 38° C and 85% relative humidity for 25 hours, and sample testing was limited to 10 minutes. The device to be tested was oriented in the filter tester so that the air and aerosol flowed from the exterior of the device toward the interior (that is, as if the wearer were inhaling, which is the same direction as when testing a respirator as a personal protective device). The challenge aerosol was generated using a 2% sodium chloride (NaCl) solution in distilled water, conditioned to 25°C and 30% relative humidity and neutralized to the Boltzmann equilibrium state. The challenge aerosol had a count median diameter of 75 nm ± 20 nm, a mass mean diameter of 260 nm and a geometric standard deviation (GSD) ≤ 1.86 (TSI 8130A specifications). The automated filter tester compares particle concentration readings from upstream and downstream light-scattering laser photometers to calculate the material filtration efficiency. An electronic pressure transducer measures the pressure difference across the material sample to indicate airflow resistance. Tests were performed with an airflow of 85 L/minute.

### Fit tests on human subjects

A convenience sample of eleven subjects (six males and five females) participated in fit testing. Subjects complied with CDC/NIOSH guidelines for facial hair styles intended for workers who wear tight-fitting respirators (CDC 2017). Fit testing was performed with a PortaCount® fit tester (model 8038, TSI) using OSHA’s modified ambient aerosol condensation nuclei counter (CNC) protocol for filtering facepiece respirators (OSHA 2019). Because only fit factors were measured and no identifiable private information was collected, the West Virginia University Office of Human Research Protections determined that Institutional Review Board approval was not required. This activity was reviewed by CDC and was conducted consistent with applicable federal law and CDC policy (see e.g., 45 C.F.R. part 46; 21 C.F.R. part 56; 42 U.S.C. §241(d), 5 U.S.C. §552a, 44 U.S.C. §3501 et seq.). All measurements were performed in a room with volume of 43.4 m^3^ (1531 ft^3^) at 19 °C (67 °F).

An NaCl aerosol generator (Model 8026, TI) was used to supplement the naturally occurring particles in the air. The particle generator was placed 10 feet away from all sampling apparatus and fit testing equipment. The generator was turned on ten minutes prior to testing to seed aerosols and turned off while a triplicate series of fit tests were performed (around 7 minutes). As the aerosol concentration in the room had started to slightly decline at this point, the generator was turned on between tests (around 7 minutes) and then turned off during subsequent tests. This process was repeated until testing for the day was completed.

To verify that suitable aerosol concentrations were present during testing, particle concentrations were measured with a condensation particle counter (CPC Model 3775, TSI) for 25 minutes beginning at 2 minutes prior to the nebulizer being initially turned on. Particle size distributions were measured for 20 minutes with a scanning mobility particle sizer (SMPS Model 3938, TSI), an aerosol particle sizer (APS Model 3321, TSI), and an electrical low-pressure impactor (ELPI+, Dakati). Each of the three particle size analyzers uses different techniques to estimate particle size and each covers different size ranges. Information on the aerosol particle size distribution and concentration are shown in the SI (Figures S5 and S6).

Each source control device was tested by three subjects; for each subject three replicate measurements were made using each of the two protocols described below. One individual administered all fit tests. Subjects were instructed in the proper donning of medical masks and respirators, and they were asked to don cloth masks in the manner that they ordinarily don similar coverings for public use. If a cloth mask was too large, it was modified using materials that would be accessible to most lay individuals: tape was used to shorten mask ear loops or head straps, and a binder clip was used to decrease the circumference of neck gaiters. These adjustments were made so that the covering was held flush to the face, but the materials were not under tension.

A fit factor was calculated by the PortaCount® software for all measurements (Janssen and McKay 2017; TSI 2015). The fit factor (FF) is calculated as:

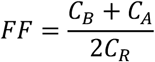

Where

C_B_ = particle concentration in the ambient sample taken before the respirator sample.

C_A_ = particle concentration in the ambient sample taken after the respirator sample.

C_R_ = particle concentration in the respirator sample.

Two particle measurement protocols were performed on all source control devices: the PortaCount® N95 Companion protocol (referred to here as N95 mode), which counts particles 0.025 – 0.06 μm in diameter; and the PortaCount® standard protocol (referred to here as all particle sizes mode), which counts particles ranging from 0.02 – 1.0 μm (TSI 2010).

### Statistical Analysis

The source control performance of each source control device was evaluated by calculating the collection efficiency, defined as:

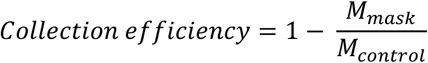

Where:

M_mask_ = total mass of the aerosol particles that passed through or around the source control device and was collected by the impactor.

M_control_ = total mass of the aerosol particles collected by the impactor while not wearing a source control device.

The aerosol masses collected by the cascade impactor during the experiments are shown in Tables S1 and S2 in the SI. To control for variations in the amount of aerosol in each experiment, a sample of each test aerosol was collected from the bellows prior to coughing or breathing and used to normalize the aerosol mass collection results for each experiment.

The source control results were compared with the filtration efficiencies, airflow resistances, and fit factor measurements by calculating the correlation coefficients (r) between the metrics. Fit factors were transformed into collection efficiencies (= 1-1/fit factor) to allow for a direct comparison with the other parameters. Because the replicate numbers were different for the different performance metrics, the correlations were calculated based on the mean results for each device. The p-value was calculated based on r and the number of types of devices that were tested (22 for all source control devices and 18 for the cloth masks alone, with the 1-layer and 2-layer gaiter tests counted separately). Correlations were considered significant if p ≤ 0.05.

## Results

### Efficacy of N95 respirators, medical masks, and cloth masks as source control devices for respiratory aerosols

Twenty-six cough experiments were performed without a source control device to measure the cough aerosol output from the source control measurement system. The cough aerosol had a geometric mean aerodynamic diameter of 1.3 µm with a GSD of 2.3 and a mean mass of 525 µg (standard deviation (SD) 65; Figure 2). Thirty-five breathing aerosol experiments were performed without a source control device to measure the exhaled aerosol output. The exhaled aerosol had a geometric mean aerodynamic diameter of 1.3 µm with a GSD of 2.3 and a mean mass of 495 µg (SD 68; Figure 2). The experiments without source control devices were used as control experiments when evaluating performance of the devices.

**Figure 2:**
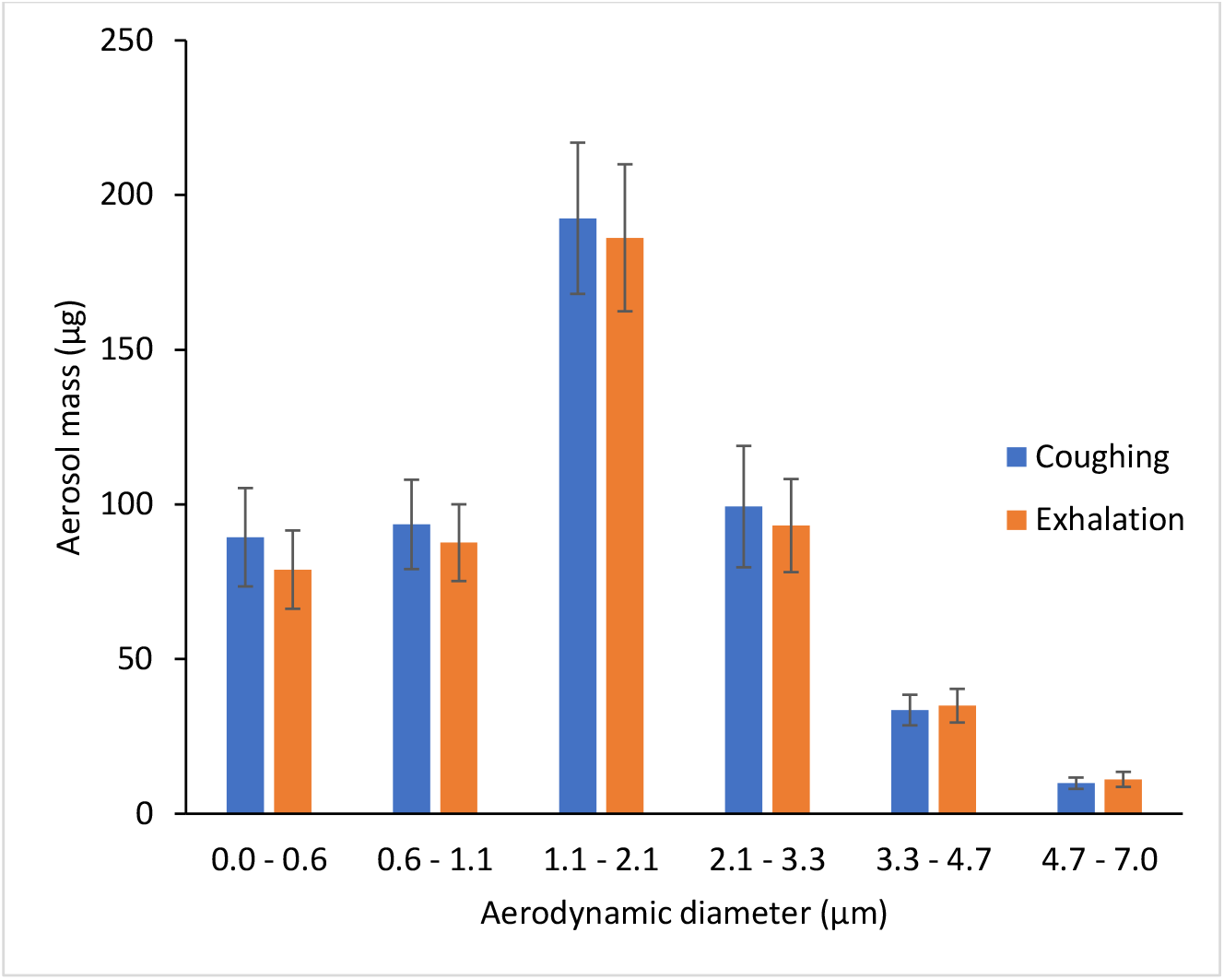
Size distribution of the cough and exhalation aerosols expelled by the source control measurement system.

The collection efficiencies of the 19 source control devices are shown in Figure 3 and Table 2. The mean collection efficiencies of the N95 respirators and the surgical mask ranged from 83% to 99% for aerosol particles during coughing and exhalation. For the procedure and cloth masks, the mean collection efficiencies ranged from 35% to 71% for coughing (except for the bandana at 17%) and 35% to 66% for exhalation. The collection efficiencies for the coughing and breathing source control experiments were reasonably well-correlated, with a correlation coefficient (r) of 0.85 (p < 0.0001; Table 3) when considering all the devices and 0.65 (p = 0.0018; Table 4) when considering only the cloth face masks, gaiters and bandanas.

**Table 2:** Collection efficiencies and fit factors for source control devices. Manikin fit factors were measured with the fit tester in N95 mode only (as described in the Discussion). Human fit factors were measured using both the N95 mode and all particle sizes mode.

| Face covering | COUGHING |  |  |  | EXHALATION |  |  |  | Filtration Efficiency (%) |  | Inhalation airflow resistance (Pa) |  | Human subject fit factor |  |  |  |
| --- | --- | --- | --- | --- | --- | --- | --- | --- | --- | --- | --- | --- | --- | --- | --- | --- |
|  | Collection efficiency (%) |  | Manikin fit factor |  | Collection efficiency (%) |  | Manikin fit factor |  |  |  |  |  | N95 mode |  | All sizes |  |
|  | mean | SD | mean | SD | mean | SD | mean | SD | mean | SD | mean | SD | mean | SD | mean | SD |
| 3M 1860 N95 respirator | 98.6% | 0.3% | 198.0 | 3.5 | 99.0% | 0.7% | 132.5 | 78.5 | 99.1% | 0.1% | 72.2 | 8.1 | 163.8 | 56.2 | 45.3 | 10.9 |
| BYD N95 respirator | 82.7% | 2.1% | 25.0 | 11.3 | 95.2% | 0.6% | 113.5 | 122.3 | 99.1% | 0.1% | 150.8 | 14.5 | 147.9 | 33.5 | 54.9 | 7.8 |
| 3M 1818 surgical mask | 86.8% | 6.3% | 26.1 | 26.7 | 91.4% | 2.6% | 27.5 | 12.0 | 92.7% | 0.6% | 48.0 | 7.8 | 78.6 | 92.0 | 9.6 | 5.8 |
| Excellent Artisan procedure mask | 56.3% | 5.8% | 7.4 | 3.4 | 42.0% | 6.7% | 6.1 | 5.5 | 79.7% | 3.6% | 108.0 | 34.3 | 3.3 | 0.7 | 1.7 | 0.5 |
| Hanes cloth mask | 51.7% | 7.1% | 1.3 | 0.1 | 44.3% | 14.0% | 2.3 | 1.1 | 18.8% | 2.7% | 64.1 | 9.5 | 1.7 | 0.5 | 1.0 | 0 |
| EnerPlex mask | 48.0% | 6.7% | 5.1 | 1.3 | 52.6% | 7.3% | 2.5 | 0.4 | 19.9% | 3.9% | 23.8 | 10.7 | 2.1 | 1.2 | 1.8 | 0.7 |
| Craft & Soul mask | 47.4% | 6.2% | 2.5 | 1.2 | 51.2% | 12.2% | 2.3 | 1.0 | 9.7% | 3.9% | 11.4 | 8.2 | 2.0 | 0.5 | 1.3 | 0.5 |
| Lefty mask | 42.3% | 8.1% | 2.4 | 0.7 | 36.3% | 9.9% | 1.9 | n/a | 20.2% | 0.8% | 96.3 | 5.9 | 1.8 | 0.7 | 1.6 | 0.5 |
| Badger dust mask | 71.1% | 3.5% | 5.0 | 0.1 | 61.8% | 5.0% | 3.2 | 0.6 | 36.0% | 9.9% | 92.1 | 1.0 | 2.8 | 1.6 | 1.7 | 0.5 |
| Debrief Me mask w filter | 43.0% | 12.0% | 4.6 | 1.8 | 63.4% | 15.3% | 3.0 | 1.5 | 30.8% | 1.5% | 154.1 | 34.9 | 2.9 | 0.9 | 2.1 | 0.8 |
| Fabrique Innovations mask | 63.2% | 6.2% | 2.1 | n/a | 65.9% | 4.4% | 2.2 | 0.4 | 14.8% | n/a | 61.7 | n/a | 1.4 | 0.7 | 1.1 | 0.3 |
| Besungo sports mask w filter | 65.7% | 6.5% | 4.8 | n/a | 59.9% | 11.9% | 3.2 | 0.3 | 97.8% | 0.3% | 142.7 | 22.9 | 2.7 | 1.7 | 1.9 | 1.4 |
| Inspire mask w filter | 66.5% | 4.5% | 4.3 | 3.0 | 63.1% | 6.4% | 2.7 | 0.5 | 27.1% | 5.1% | 149.8 | 85.4 | 4.0 | 1.4 | 2.4 | 0.5 |
| FKGIONG gaiter 1-layer | 48.1% | 6.7% | 1.7 | 0.5 | 42.9% | 5.5% | 1.5 | 0.2 | 3.3% | 4.3% | 13.1 | 8.0 | 1.1 | 0.3 | 1.0 | 0 |
| FKGIONG gaiter 2-layers | 60.5% | 6.7% | 1.9 | 0.4 | 51.8% | 8.6% | 1.7 | n/a | 7.9% | 0.5% | 23.8 | 8.4 | 1.0 | 0 | 1.4 | 0.5 |
| AXBXCX gaiter 1-layer | 35.4% | 9.5% | 1.7 | 0.1 | 35.3% | 10.3% | 1.4 | 0.1 | 2.7% | 1.1% | 14.4 | 5.7 | 1.3 | 0.7 | 1.0 | 0 |
| AXBXCX gaiter 2-layers | 48.3% | 6.3% | 1.8 | 0.1 | 43.9% | 5.6% | 1.9 | 0.5 | 3.6% | 0.3% | 29.4 | 4.5 | 1.3 | 0.5 | 1.3 | 0.5 |
| Retro gaiter 1-layer | 48.7% | 10.5% | 2.6 | n/a | 55.8% | 6.6% | 2.5 | 0.6 | 2.2% | n/a | 40.2 | n/a | 1.6 | 0.7 | 1.3 | 0.5 |
| Retro gaiter 2-layers | 64.9% | 4.2% | 2.3 | 0.4 | 60.0% | 5.5% | 2.1 | 0.4 | 8.6% | n/a | 69.5 | n/a | 1.7 | 1.0 | 1.7 | 0.5 |
| Givon fleece gaiter 1-layer | 53.5% | 6.3% | 2.1 | 0.5 | 51.4% | 7.0% | 2.3 | 0.8 | 8.4% | 0.8% | 19.3 | 7.4 | 1.0 | 0 | 1.3 | 0.5 |
| L&M Bandana 2-layers | 17.1% | 25.0% | 1.3 | 0.2 | 46.5% | 8.1% | 1.6 | 0.1 | 1.4% | 0.4% | 5.2 | 8.2 | 1.1 | 0.3 | 1.0 | 0 |
| Underwear Bandana/gaiter 1-layer | 40.8% | 7.5% | 6.9 | 7.3 | 37.3% | 10.2% | 1.4 | 0.1 | 3.5% | 1.1% | 19.3 | 8.2 | 1.1 | 0.7 | 1.0 | 0 |

**Table 3:** Correlations between different performance metrics for all source control devices. R indicates the correlation coefficient while r^2^ is the coefficient of determination.

| *Correlation parameter #1* | *Correlation parameter #2* | *# points* | *r* | *r<sup>2</sup>* | *p-value* |
| --- | --- | --- | --- | --- | --- |
| Cough aerosol collection efficiency | Exhaled aerosol collection efficiency | 22 | 0.8453 | 71% | <0.0001 |
|  | Manikin fit before coughing | 22 | 0.5911 | 35% | 0.0019 |
|  | Filtration efficiency of material | 22 | 0.7428 | 55% | <0.0001 |
|  | Airflow resistance of material | 22 | 0.4280 | 18% | 0.0235 |
|  | Human subject fit (N95 mode) | 22 | 0.7120 | 51% | 0.0001 |
|  | Human subject fit (all particle sizes) | 22 | 0.8054 | 65% | <0.0001 |
| Exhaled aerosol collection efficiency | Manikin fit before breathing | 22 | 0.8863 | 79% | <0.0001 |
|  | Filtration efficiency of material | 22 | 0.7199 | 52% | 0.0001 |
|  | Airflow resistance of material | 22 | 0.4140 | 17% | 0.0277 |
|  | Human subject fit (N95 mode) | 22 | 0.7528 | 57% | <0.0001 |
|  | Human subject fit (all particle sizes) | 22 | 0.8585 | 74% | <0.0001 |
| Manikin fit before coughing | Manikin fit before breathing | 22 | 0.6389 | 41% | 0.0007 |
| Filtration efficiency of material | Airflow resistance of material | 22 | 0.6350 | 40% | 0.0007 |

**Table 4:** Correlations between different performance metrics for the cloth face masks, gaiters and bandanas. R indicates the correlation coefficient while r^2^ is the coefficient of determination.

| *Correlation parameter #1* | *Correlation parameter #2* | *# points* | *r* | *r<sup>2</sup>* | *p-value* |
| --- | --- | --- | --- | --- | --- |
| Cough aerosol collection efficiency | Exhaled aerosol collection efficiency | 18 | 0.6487 | 42% | 0.0018 |
|  | Manikin fit before coughing | 18 | 0.3140 | 10% | 0.1022 |
|  | Filtration efficiency of material | 18 | 0.4637 | 21% | 0.0263 |
|  | Airflow resistance of material | 18 | 0.4802 | 23% | 0.0219 |
|  | Human subject fit (N95 mode) | 18 | 0.4133 | 17% | 0.0441 |
|  | Human subject fit (all particle sizes) | 18 | 0.5352 | 29% | 0.0110 |
| Exhaled aerosol collection efficiency | Manikin fit before breathing | 18 | 0.6596 | 44% | 0.0014 |
|  | Filtration efficiency of material | 18 | 0.4352 | 19% | 0.0355 |
|  | Airflow resistance of material | 18 | 0.5697 | 32% | 0.0068 |
|  | Human subject fit (N95 mode) | 18 | 0.5390 | 29% | 0.0105 |
|  | Human subject fit (all particle sizes) | 18 | 0.6233 | 39% | 0.0029 |
| Manikin fit before coughing | Manikin fit before breathing | 18 | 0.3587 | 13% | 0.0719 |
| Filtration efficiency of material | Airflow resistance of material | 18 | 0.7185 | 52% | 0.0004 |

**Figure 3:**
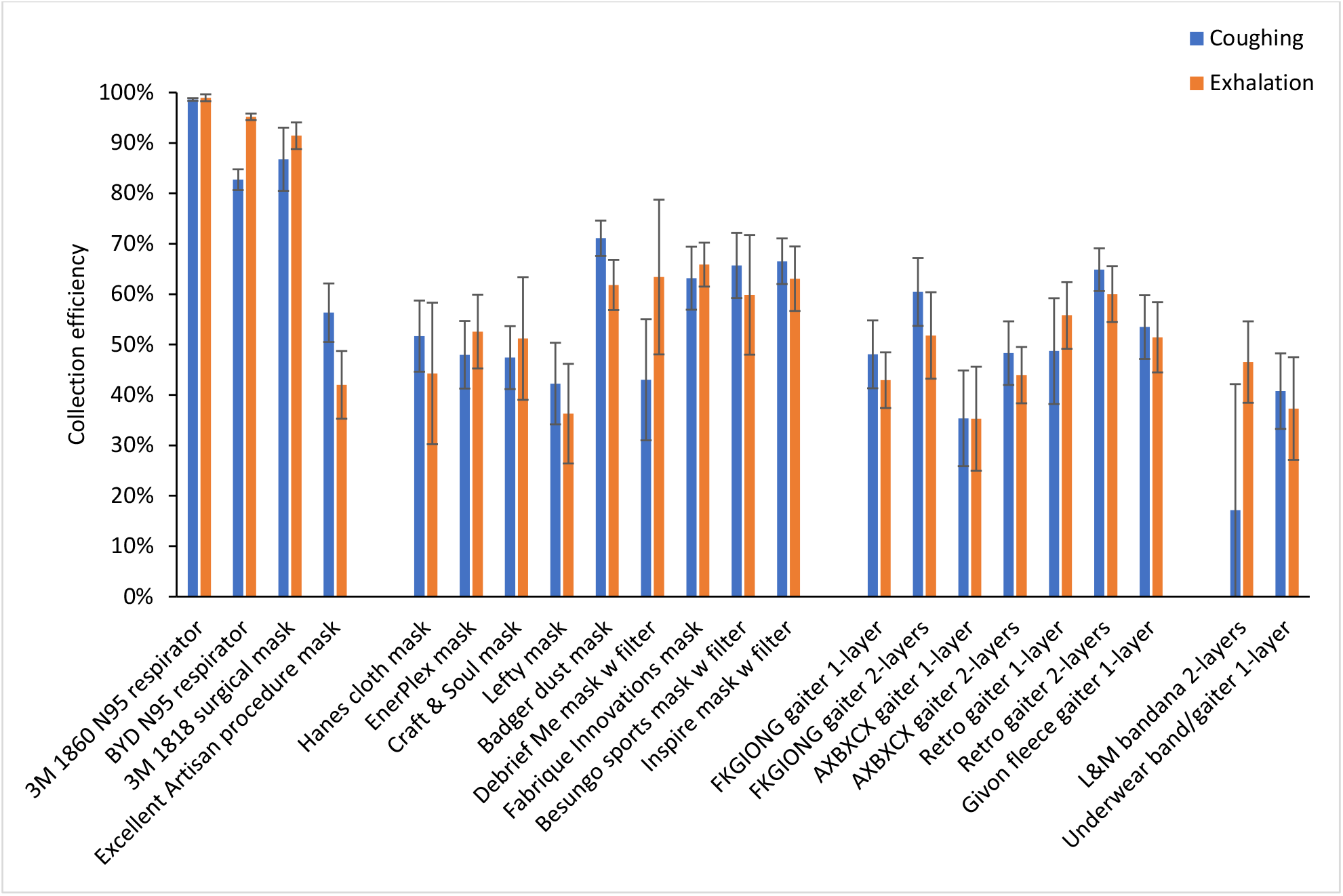
Collection efficiencies of N95 respirators, medical masks, cloth face masks, neck gaiters and bandanas during coughing and exhalation aerosol tests with the respiratory aerosol source control measurement system. Each bar is the mean of 4 or 6 experiments. Error bars show the standard deviation.

### Filtration efficiencies and inhalation airflow resistance

The mean material filtration efficiencies were >99% for the respirators, 80% to 93% for the medical masks, and 1.4% to 36% for the cloth face masks, neck gaiters, and bandanas, except for the Besungo sports mask at 98% (Figure 4 and Table 2). The inhalation airflow resistances were from 5.2 to 154 Pa (Table 2). When comparing the data for all devices, the filtration efficiencies showed a good correlation with cough aerosol collection efficiency (r = 0.74, p < 0.0001) and exhaled aerosol collection efficiency (r = 0.72, p = 0.0001; Table 3). For the group of cloth masks, the correlations were not as strong; the cough aerosol collection efficiency and filtration efficiency had an r of 0.46 (p = 0.0263) while the exhaled aerosol collection efficiency and filtration efficiency had an r of 0.44 (p = 0.0355; Table 4). The airflow resistance did not correlate as well as the filtration efficiency when all devices were considered (r = 0.43 and p = 0.0235 for coughing; r = 0.41 and p = 0.0277 for exhalation). However, when examining the cloth masks alone, the airflow resistance was slightly better correlated than filtration efficiency (r = 0.48 and p = 0.0219 for coughing; r = 0.57 and p = 0.0068 for exhalation).

**Figure 4:**
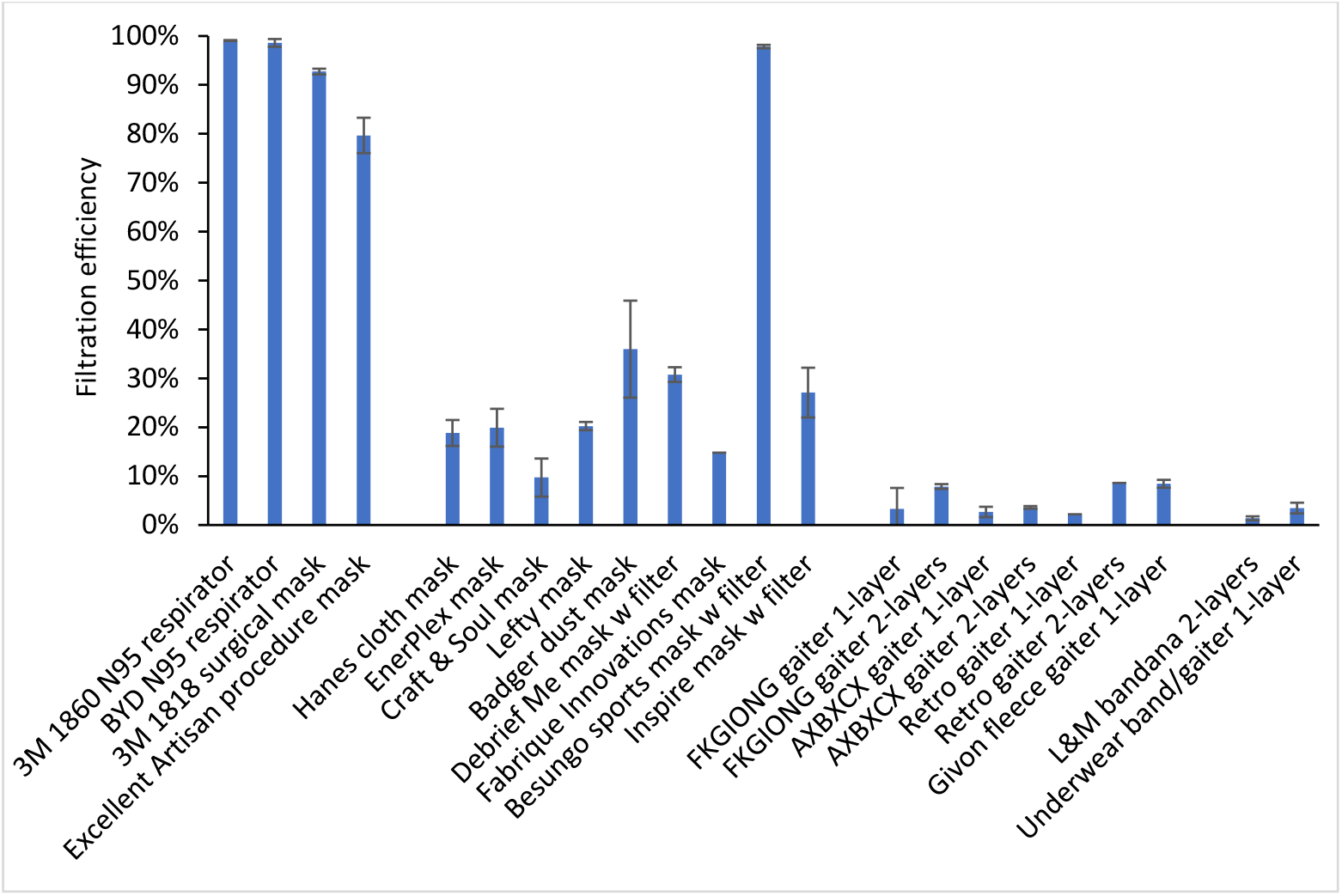
Filtration efficiencies for the source control devices used in the study. Three samples of each device were tested, except for the 3M 1860 N95 respirator and Hanes cloth mask (10 samples each) and the Fabrique Innovations mask, Retro gaiter 1-layer, and Retro gaiter 2-layers (one sample each). Error bars show the standard deviation.

### Human fit tests

The fit factors measured on the human test subjects are shown in Table 2 and Figure 5. The mean fit factors for the two N95 respirators were 147.9 and 163.8 when using the N95 mode on the fit tester and 54.9 and 45.3 with the all particle sizes mode. The surgical mask provided a mean N95 mode fit factor of 78.6 and a much lower mean fit factor of 9.6 in all particle sizes mode. The mean N95 mode fit factor of the reusable cloth face masks ranged from 1.4 to 4.0, and the all sizes mode mean fit factors ranged from 1.0 to 2.4. Neck gaiters and bandanas demonstrated N95 mode fit factors ranging from 1.0 to 1.7 and all particle size mode mean fit factors from 1.0 to 1.7. The performance of cloth masks sometimes varied considerably when worn by different subjects.

**Figure 5:**
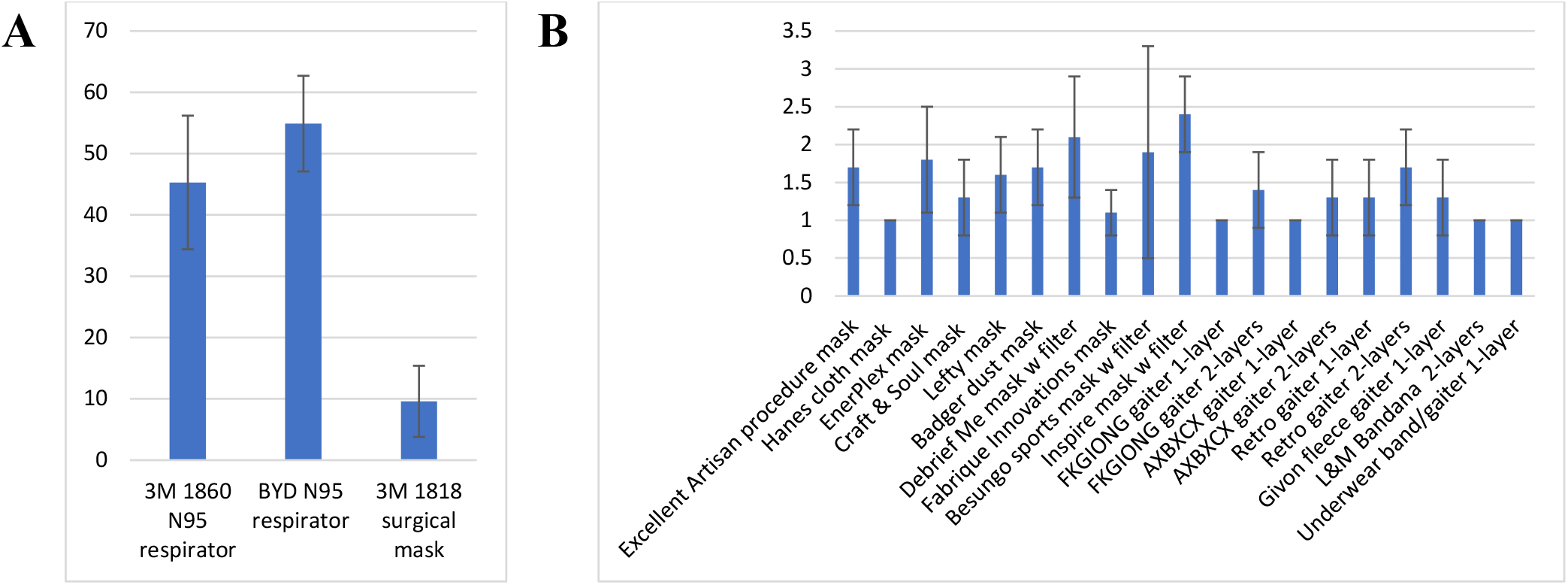
Fit factors measured with human subjects for A) N95 respirators and surgical mask and B) Procedure mask, cloth face masks, gaiters and bandanas. The data was collected with the fit tester measuring all particle sizes. The plot shows the means and standard deviations.

The cough and exhalation aerosol collection efficiencies for all devices were well correlated with the fit factors when measured using the all particle sizes mode (r= 0.81 and p < 0.0001 for coughing, r = 0.86 and p < 0.0001 for exhalation, Table 3). When looking at the results for the cloth masks, gaiters, and bandanas in the all sizes mode, the correlations were not as strong, with r = 0.54 for coughing (p = 0.0110; Table 4), and r =0.62 for exhalation (p = 0.0029; Table 4). In all cases, the fit factors were significantly correlated with the cough and exhaled aerosol collection efficiencies, and the correlation coefficients were higher when the fit tester was in the all particle sizes mode than when the tester was in N95 mode.

### Manikin fit tests

The manikin fit factor measurements found using the source control measurement system before the cough tests were 25 to 198 for the N95 respirators and the surgical mask and 1.3 to 7.4 for the procedure mask and cloth masks (Table 2). Before the breathing tests, the manikin fit factors were 28 to 133 for the N95 respirators and the surgical mask and 1.4 to 6.1 for the procedure mask and cloth masks. The cough aerosol collection efficiencies and the pre-cough manikin fit factors were somewhat correlated, with an r of 0.59 (p = 0.0019) for all devices (Table 3 and Figure 6), but an r of only 0.31 (p = 0.1022) for the cloth masks (Table 4). The correlation was better between the exhaled aerosol collection efficiency and pre-breathing manikin fit factors, with an r of 0.89 (p < 0.0001) for all devices (Table 3 and Figure 7) and 0.66 (p = 0.0014) for the cloth masks (Table 4).

**Figure 6:**
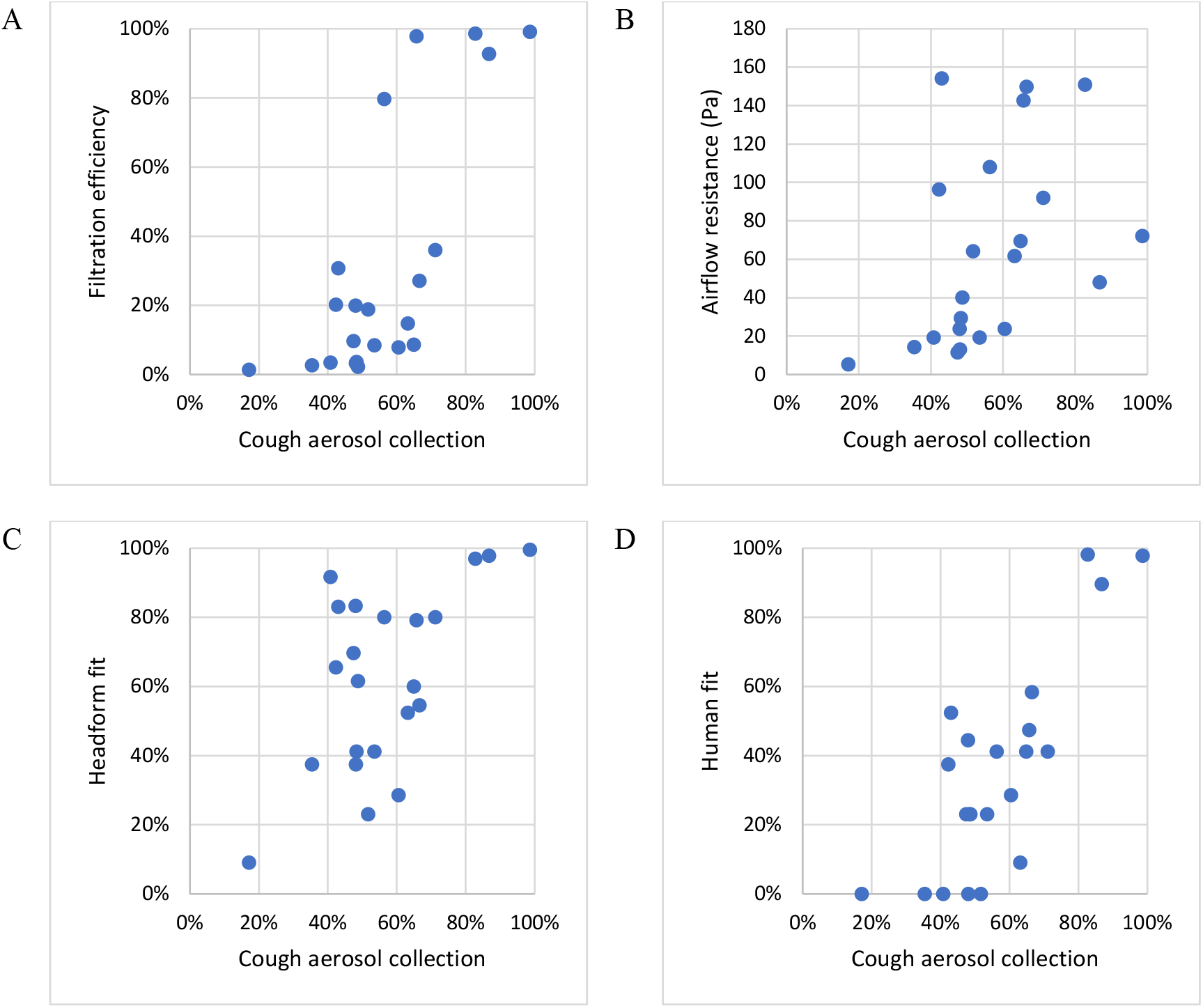
Comparison of collection efficiencies for aerosols from coughs to (A) Filtration efficiency of construction material; (B) Airflow resistance; (C) Fit on manikin headform; and (D) Fit on human test subjects (using all particle sizes mode). Each dot corresponds to the mean result for one type of source control device.

**Figure 7:**
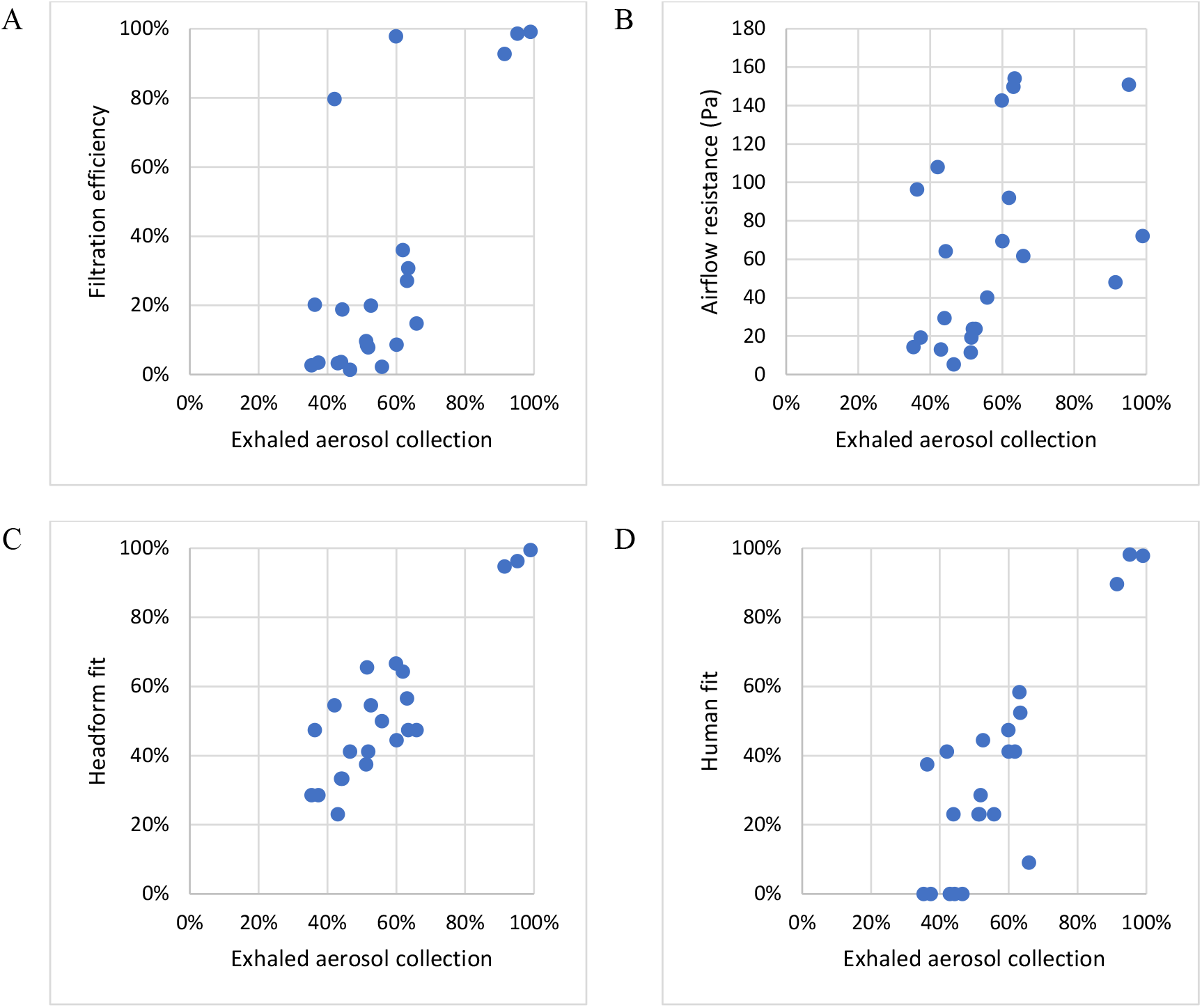
Comparison of collection efficiencies for exhaled aerosols from simulated breathing to (A) Filtration efficiency of construction material; (B) Airflow resistance; (C) Fit on manikin headform; and (D) Fit on human test subjects (using all particle sizes mode). Each dot shows the mean result for one type of source control device.

## Discussion

The COVID-19 pandemic has resulted in considerable interest in the performance characteristics of masks and respirators as a means of reducing the person-to-person transmission of SARS-CoV-2. Some studies have looked at the emission of particles from human volunteers wearing different face coverings, which has the advantage of directly examining the expulsion of aerosols into the environment by people (Asadi et al. 2020; Davies et al. 2013; Leung et al. 2020; Li et al. 2020). However, the collection and measurement of human-generated respiratory aerosols is challenging and can be hazardous when the subject has a contagious respiratory infection. In addition, the amount and size distribution of aerosol particles produced by people during coughing, breathing and other respiratory activities varies tremendously from person to person and even for a particular person over time, which makes it difficult to directly compare results (Asadi et al. 2019; Fennelly 2020; Gralton et al. 2011; Lindsley et al. 2012). Consequently, lab-based surrogate techniques are more frequently used to study source control devices. The most commonly reported methods have been tests of filtration efficiencies and fit factors (Clapp et al. 2020; Guha et al. 2021; Konda et al. 2020; Wang et al. 2020; Zhao et al. 2020). Other studies have used manikins in a chamber or room to assess the efficacy of medical masks and cloth masks both as source control devices and as personal protective equipment (Pan et al. 2020; Patel et al. 2016; Rothamer et al. 2021). Experimental data have also been used to develop computational fluid dynamics models of medical and cloth masks performance (Dbouk and Drikakis 2020; Mittal et al. 2020). These studies have provided valuable information about the properties of masks and respirators. However, it is unclear how the various metrics used in these studies are related to source control performance.

The source control measurement system used in this study allows for a direct quantitative comparison of the ability of different types of source control devices to block the expulsion of simulated cough and exhaled breath aerosol particles into the environment (Lindsley et al. 2021). However, the system is complex and requires expertise to operate, and it is neither commercially sold nor easily constructed. In contrast, filtration measurement systems and respirator fit testers are widely available. If a standard methodology could be developed to gauge source control performance using filtration testing and/or fit testing, it could be rapidly expanded and adopted by manufacturers and public health entities.

The filtration efficiency of a mask or respirator is a measurement of the ability of the construction material to remove aerosol particles from an airstream traveling through the fabric. Filtration efficiency tests typically use aerosol particles at or near the most-penetrating aerosol particle size, which is around 300 nm for an uncharged filter at low air velocities but shifts to smaller sizes at higher velocities and when the filter and particles are electrostatically charged (Martin and Moyer 2000; Rengasamy et al. 2013; Rengasamy et al. 2017). Although the SARS-CoV-2 virus is about 100 nm in diameter, contagious humans do not shed bare viral particles. Instead, they expel aerosols and droplets of respiratory fluids that contain respiratory virus, and the size of these virus-laden aerosols and droplets can range from hundreds of nanometers to visible droplets of 1 mm or more (Fennelly 2020; Gralton et al. 2011). The mechanisms by which filters collect aerosol particles are strongly dependent upon the size of the particles; for example, large particles are more likely to be collected by impaction or interception while small particles are more likely to be collected by diffusion (Lindsley 2016). Thus, the collection efficiencies for 100-300 nm particles may not predict the performance for larger particles (Drewnick et al. 2020). In our experiments, the coughing and exhalation source control collection efficiencies increased as the particle size increased (Table S1 and S2 in the SI) and in most cases were much higher than the filtration efficiencies, likely because our coughing and exhalation test aerosols were larger than the 75 nm aerosol used for filtration tests.

The fit factor is normally used to determine how well a respiratory protective device fits the wearer by measuring the degree to which aerosol particles can enter through gaps between the wearer’s face and the respirator (face seal leaks). It is determined by placing the device on a person or on a manikin headform and measuring the ratio of the aerosol concentration outside the respirator to the aerosol concentration inside the respirator (Janssen and McKay 2017). For example, a fit factor of 10 means that the ambient aerosol concentration is 10 times higher than the concentration inside the respirator. Fit factor measurements are not intended to test the filtration efficiency of the device itself. In fact, the calculation of the fit factor for a respirator assumes that any ambient aerosol particles passing through respirator material are filtered out and that any particles detected inside the respirator are due to face seal leaks, not penetration of particles through the filter material (Halvorsen 1998). For example, a fit factor of 10 is interpreted as indicating that 90% of the air inside the respirator has passed through the respirator filtration media (with aerosol particles being completely removed) and that 10% of the air bypassed the media and entered through face seal leaks.

For high efficiency filters such as P100 respirator filters, which filter out 99.97% of airborne particles, a respirator fit test is performed by measuring the concentrations of aerosol particles of all sizes inside and outside the respirator. However, N95 respirators can allow up to 5% of aerosol particles of the most penetrating size to pass through the filtration media. These particles would reduce the apparent fit factor if they were included in the calculation. Thus, when fit testing N95 respirators, the respirator fit tester used in our experiments has a size classifier to count only negatively charged aerosol particles near 55 nm in size (referred to here as N95 mode). For an N95 respirator, these charged 55 nm particles are almost entirely filtered out by the filtration media, and thus any particles detected inside the respirator can then be assumed to have entered through face seal leaks and not through the respirator (Halvorsen 1998; Han and Prell 2010).

It is important to note, however, that this assumption is not correct when using the fit tester with cloth masks. As can be seen in Table 2, the filtration efficiencies of most cloth masks are much lower than those of respirators, and ambient particles therefore can penetrate more easily through the cloth material. Thus, the fit factor for a cloth mask is not a true measurement of face seal leakage alone; instead, it represents a combination of face seal leakage and particle penetration through the mask material.

Our findings suggest that the results from filtration efficiency and fit factor tests are significantly correlated with measurements made using our source control measurement system. However, our findings also showed that those relationships generally were not very strong. The coefficient of determination (r^2^) indicates how much the variation in one parameter can be explained by the variation in a second parameter. For example, an r^2^ value of 67% would mean that two-thirds of the changes in one parameter can be explained by changes in the other parameter, and that the remaining one-third of the variation is due to other factors or experimental noise. When comparing the source control performance for coughed and exhaled aerosols to the other performance metrics for all the devices, r^2^ ranged from 17% to 79% and was statistically significant in all cases (Table 3). When looking only at the cloth masks in our tests, r^2^ ranged from 10% to 44% (Table 4), which means that even the best metrics explained less than half of the variation in source control performance. For the cloth masks, all but one of the other performance metrics were significantly correlated with the cough and exhaled aerosol collection efficiencies, which suggests that these other measures could be useful as part of a method for testing the performance of cloth masks. However, none of the metrics were strong predictors of source control performance, and no metric was clearly superior to the others.

The limited correlations between the different test methods in our study likely have several explanations. First, filtration efficiency and airflow resistance measurements do not account for how well the devices fit the wearer or for face seal leaks around the edges of the device. Thus, for example, the Artisan procedure mask had an 80% filtration efficiency but a 56% collection efficiency for cough aerosols and a 42% collection efficiency for exhaled aerosols. The most likely explanation is that this mask’s material is an efficient filter but that aerosol particles were able to travel around the mask through face seal leaks. On the other hand, the Hanes Defender cloth mask had only a 19% filtration efficiency but had similar aerosol collection efficiencies of 52% for coughs and 44% for exhalations. In this case, the cloth mask was not an effective filter for the 75 nm aerosol used in filtration testing, but the mask fit the manikin headform more tightly. Thus, much of the aerosol then flowed through the mask rather than around it during coughing and breathing tests and the larger aerosol particles that were used in the coughing and breathing tests were filtered out more effectively than the 75 nm particles used in filtration testing. The results presented here are consistent with a previous study by our group in which knotting the Artisan procedure mask to improve the fit increased the cough aerosol collection efficiency to 77%, while wearing a Hanes Defender cloth mask on top of an Artisan procedure mask increased the cough collection efficiency to 85% (Brooks et al. 2021). Together, these results also support the recommendation by Gandhi and Marr that members of the public wear a high-quality mask that fits tightly or wear a tight-fitting cloth mask over a surgical mask in order to reduce face seal leaks and improve the source control performance and protection offered by the masks (Gandhi and Marr 2021).

Fit tests are designed to measure the effects of face seal leaks but have their own limitations when applied to cloth masks. As noted above, fit factor measurements for cloth masks reflect some combination of particle penetration through the media and face seal leakage which will likely vary from mask to mask. Thus, it is neither clear how to interpret fit factor measurements for cloth masks nor is it clear how this relates to the effectiveness of the mask as a source control device. In addition, fit test results can vary greatly from person to person and even somewhat for the same person during repeated tests (Lawrence et al. 2006). Similarly, a comparison of fit tests between humans and a pliable skin manikin headform found significant differences (Bergman et al. 2015). Much of this discrepancy is likely due to facial variations and differences in how the mask is placed on the person or manikin headform. It is possible to shift, stretch, tighten, loosen, or adjust the masks in many ways, and small differences in how the mask is worn may have substantial effects on the fit test results. For example, in tests with three subjects using the Artisan procedure mask, we found that tightening the mask against the face by using silicone ear loop adjusters increased the mean fit factor from 1.7 to 4.0 (Figure 8). In our tests of cloth masks, the r^2^ value for the manikin headform fit tests done before coughing experiments and before exhalation experiments was only 13%, suggesting that differences were occurring even when the same model of mask was being placed on the same headform.

**Figure 8:**
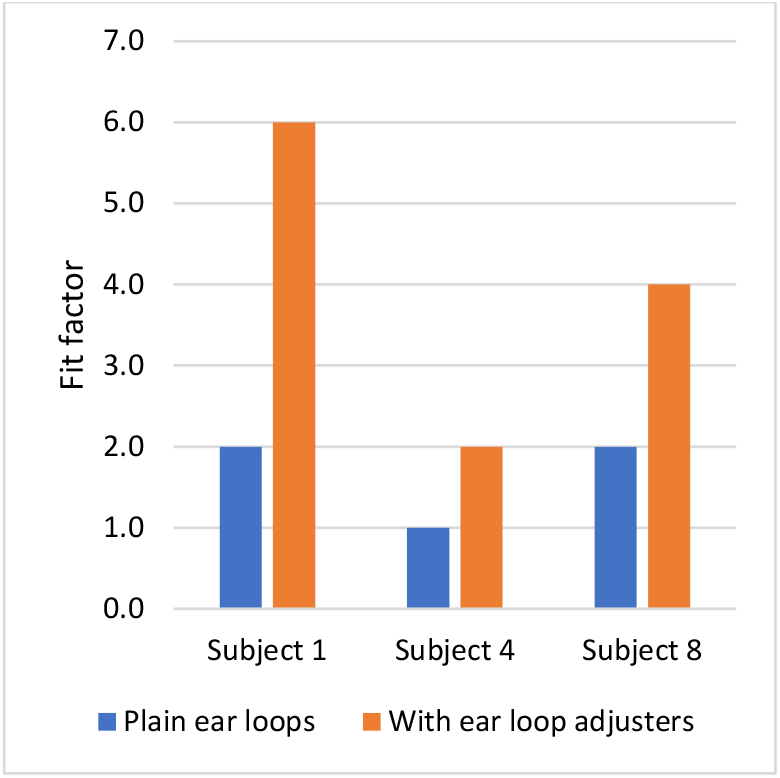
Effect on mask fit factor of adding ear loop adjusters to tighten the mask fit. Subjects were wearing the Excellent Artisan procedure mask. Tests were conducted with the fit tester in all particle sizes mode. Each bar is the mean of three trials. All standard deviations were zero.

Finally, our study has several limitations. The source control measurement system produced a test aerosol with particles ≤ 7 µm in diameter, which is the size range of aerosol particles most likely to remain airborne and most difficult to block with source control devices. However, humans expel aerosol particles in a much broader range of sizes, particularly when coughing. We used a single cough flowrate profile and a single breathing ventilation rate for our studies; these parameters can vary greatly from person to person under different physiological conditions. Some internal losses of the test aerosol particles likely occurred due to settling or impaction on the surfaces of the collection chamber, which may affect the estimates of the collection efficiencies. We used representative examples of different types of source control devices, but many such devices are available with a wide range of shapes and compositions, which would be expected to affect their individual performance. Lastly, because of the need to rapidly produce results in response to the COVID-19 pandemic, we only tested 2-3 samples of each source control device (each sample was used in two consecutive tests). By comparison, for example, the NIOSH procedure for evaluating N95 respirators calls for 20 samples to be tested (NIOSH 2019).

Current evidence indicates that masks like those tested in our experiments can substantially decrease the amount of respiratory aerosols released by the wearer, and also help reduce what the wearer breathes in (CDC 2020c). Both effects vary depending upon the material and construction of the mask, as well as how it is worn. In addition to consistent and correct mask use when in the company of others, other measures such as physical separation are important, particularly during brief exposures. In a room where someone sneezes, being six feet or further away is better than being closer. However, with prolonged exposure in the same space for more than a few minutes, the benefit of distance fades as exhaled respiratory aerosols drift, mix, and accumulate in the enclosed air space. Optimizing ventilation, air filtration, and the introduction of fresh air can help counter this effect, but at every distance, correct mask use reduces the risk for everyone.

## Conclusions

The ongoing COVID-19 pandemic has created an urgent need for simple methods to evaluate cloth masks as respiratory aerosol source control devices and to produce meaningful performance metrics that can be used to determine the effectiveness of different materials and designs. Our results suggest that test methods such as filtration efficiency testing, airflow resistance testing, and fit factor measurements on manikin headforms or human subjects have potential as ways to estimate the performance of masks as source control devices for respiratory aerosols. However, more research and improvements in the test methodologies are needed before such methods can be reliably implemented. Until the factors controlling the performance of source control devices are better understood and better testing methodologies are developed, results from test methods such as filtration efficiency testing, airflow resistance testing, and fit factor measurements should be interpreted cautiously when used to evaluate source control devices for respiratory aerosols.

## Data Availability

The experimental data is provided in the manuscript tables and the supplemental information. More detailed data is available upon request.

## Acknowledgments

We would like to thank Dr. Michael J. Beach, Dr. Michael Bell and Dr. John T. Brooks of the CDC COVID-19 Distancing/Masking Tiger Team for their advice and support. We also would like to thank the NIOSH Morgantown maintenance, security, warehouse and housekeeping departments for their assistance and dedication during the ongoing pandemic. The findings and conclusions in this report are those of the authors and do not necessarily represent the official position of the National Institute for Occupational Safety and Health (NIOSH), US Centers for Disease Control and Prevention (CDC). Mention of any company or product does not constitute endorsement by NIOSH or CDC.

## Declaration of Interests Statement

The authors declare no competing interests.

## Funding

This work was supported by the CDC and by the National Institutes of Health under Grants NIH R01 ES015022 (TRN) and NIH U54 GM104942 to the West Virginia University researchers.

## Notes

### Competing Interest Statement

The authors have declared no competing interest.

### Clinical Trial

This study was not a clinical trial

### Author Declarations

Because only fit factors were measured on human subjects and no identifiable private information was collected, the West Virginia University Office of Human Research Protections determined that Institutional Review Board approval was not required. This activity was reviewed by CDC and was conducted consistent with applicable federal law and CDC policy (see e.g., 45 C.F.R. part 46; 21 C.F.R. part 56; 42 U.S.C. 241(d), 5 U.S.C. 552a, 44 U.S.C. 3501 et seq.).

