## Supplemental materials for "A comparison of performance metrics for cloth face masks as source control devices for simulated cough and exhalation aerosols"

### A comparison of performance metrics for face coverings as aerosol source control devices during coughing and breathing

### Online Supplementary Information

| | | |
|---|---|---|
| 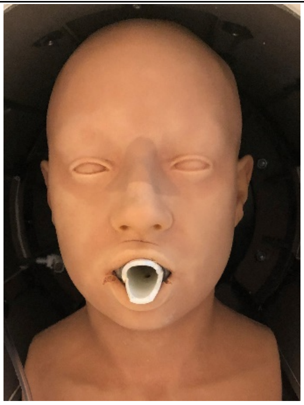<br>Pliable skin head form | 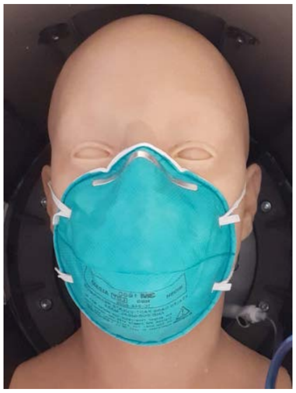<br>3M 1860 N95 respirator | 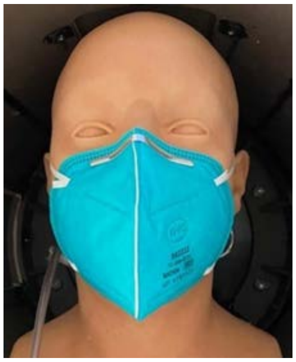<br>BYD N95 respirator |
| 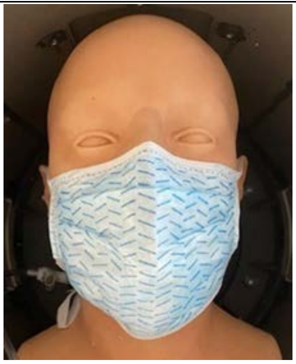<br>3M 1818 surgical mask | 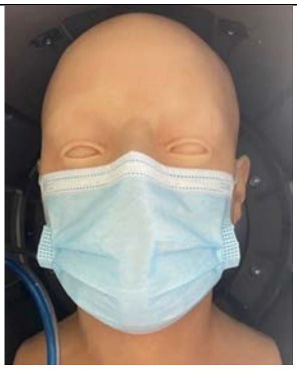<br>Excellent Artisan procedure mask | |

Figure S1. Medical masks and respirators tested as source control devices.

| | | | |
|---|---|---|---|
| 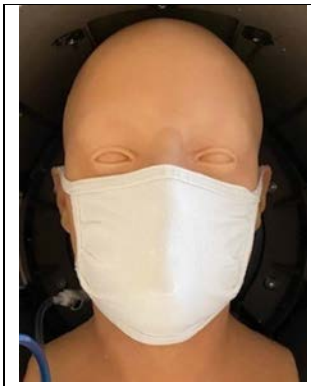 Hanes cloth mask | 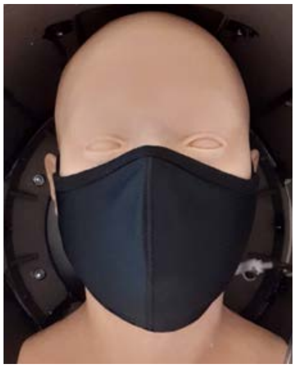 EnerPlex mask | 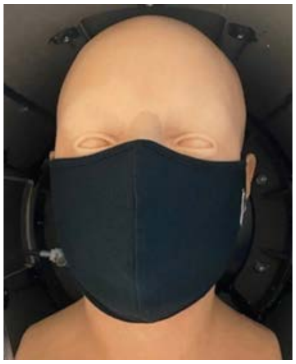 Craft & Soul mask | 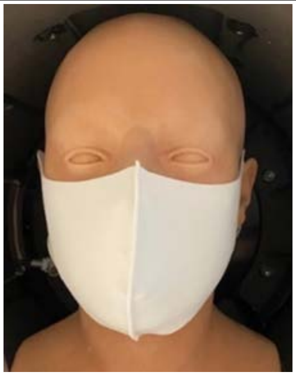 Lefty mask |
| 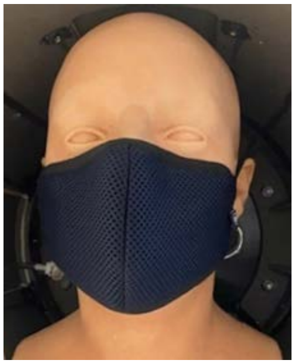 Badger dust mask | 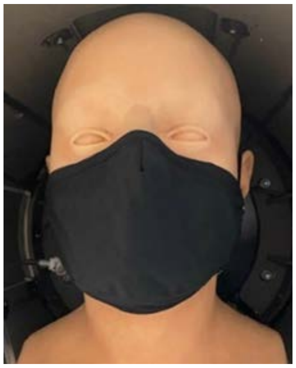 Debrief Me mask with filter | 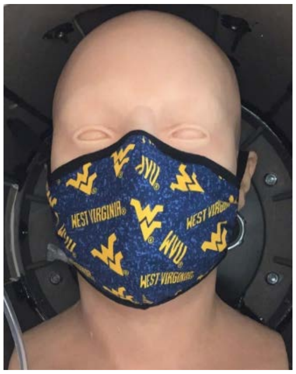  Fabrique Innovations mask | 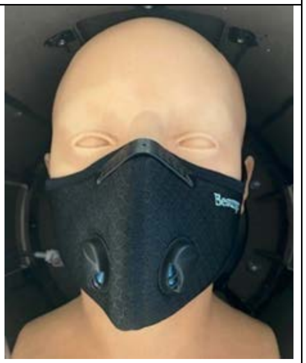 Besungo sports mask with filter and exhalation valves |
| 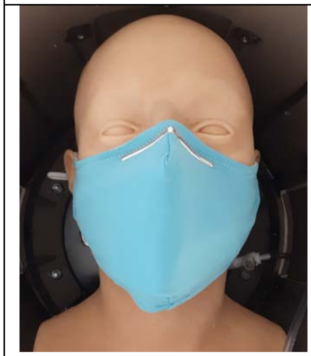 Inspire mask with filter | | | |

Figure S2. Reusable cloth face masks tested as source control devices.

| | | | |
|---|---|---|---|
| 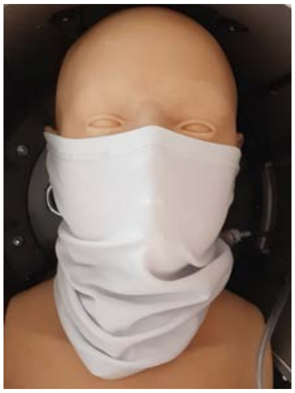 FKGIONG gaiter 1-layer | 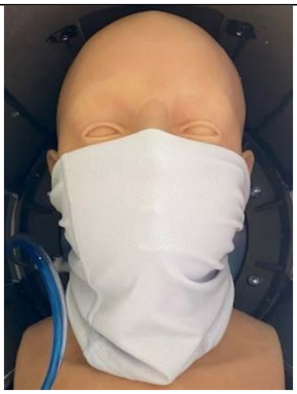 FKGIONG gaiter 2-layers | 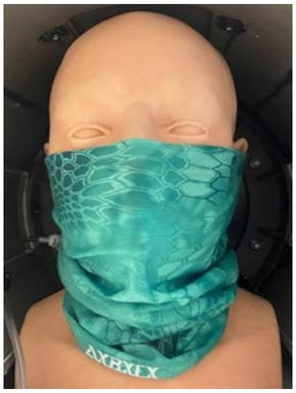 AXBXCX gaiter 1-layer | 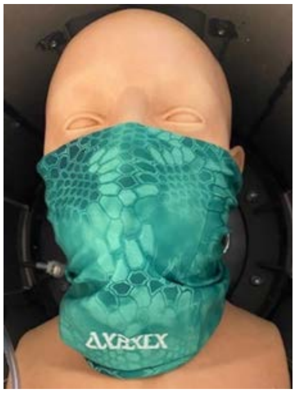 AXBXCX gaiter 2-layers |
| 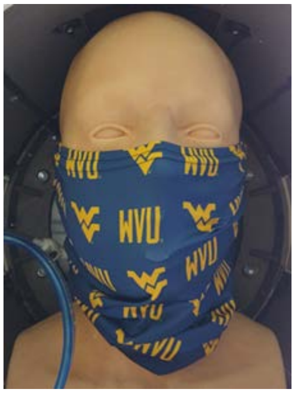 Retro gaiter 1-layer | 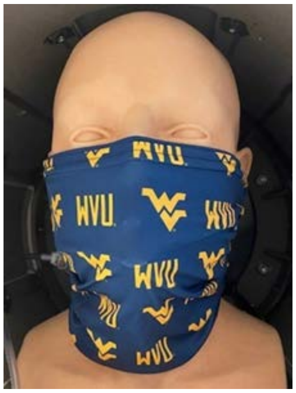 Retro gaiter 2-layers | 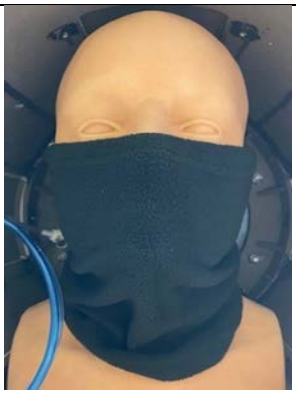 Givon fleece gaiter 1-layer | |
| 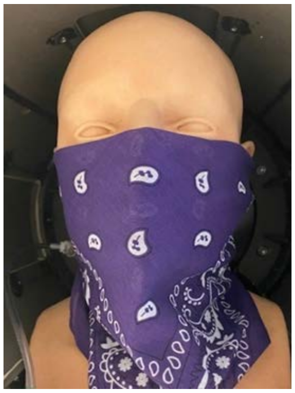 L&M Bandana 2-layers | 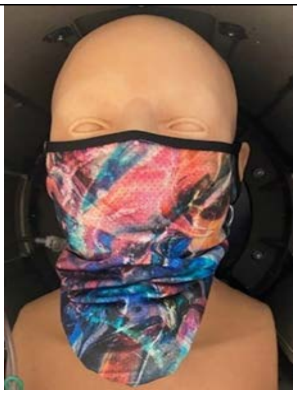 Underwear bandana/ gaiter 1-layer | | |

Figure S3. Neck gaiters and bandanas tested as source control devices.

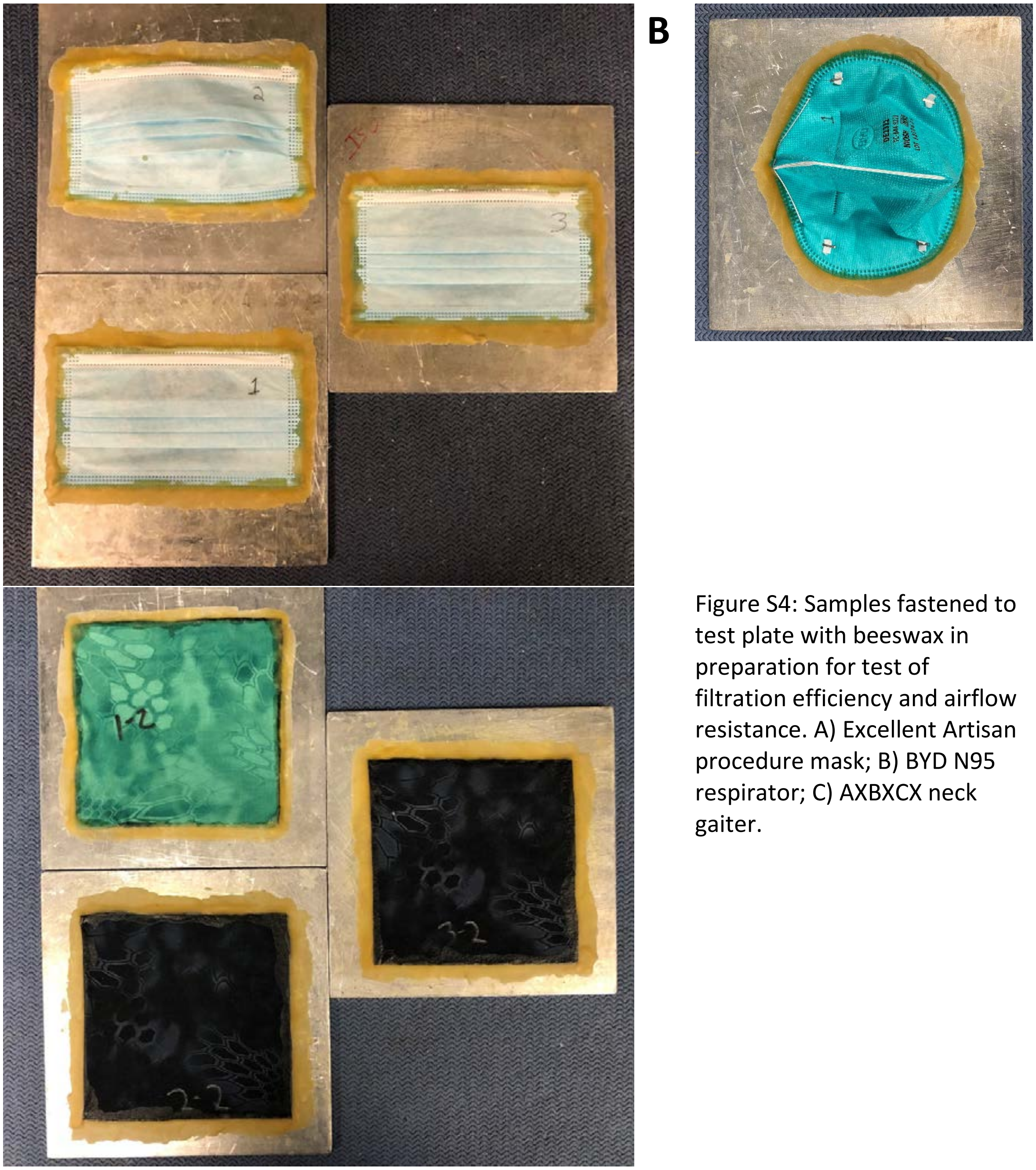


Figure S4: Samples fastened to test plate with beeswax in preparation for test of filtration efficiency and airflow resistance. A) Excellent Artisan procedure mask; B) BYD N95 respirator; C) AXBXCX neck gaiter.

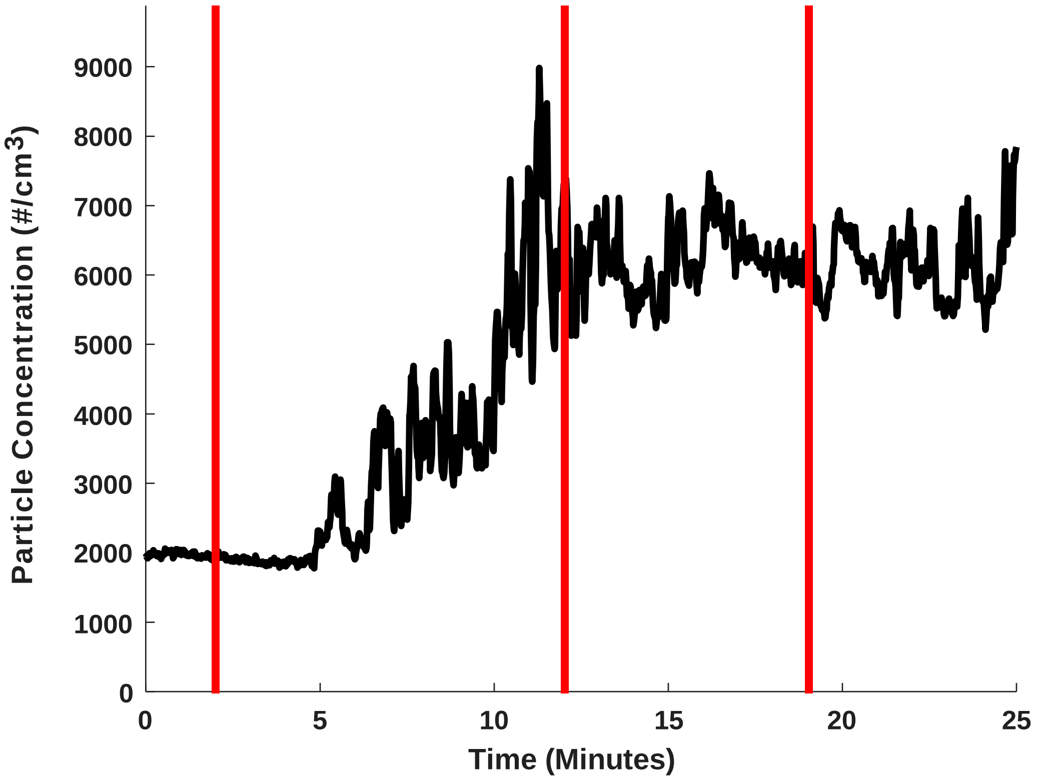


Figure S5: Saline particle concentration measured from the room during testing with a condensation particle counter as the particle generator is turned on (first red line), turned off (second red line) and turned on again (third red line). Intermittently switching the nebulizer on and off permitted an acceptable concentration of aerosols (between 1,000-30,000 particles/cm$^3$) (TSI 2015) to be present in the room during fit testing. After the room was initially filled with the saline particles, the concentration in the room typically ranged between 5000-8000 particles/cm$^3$ during fit testing.

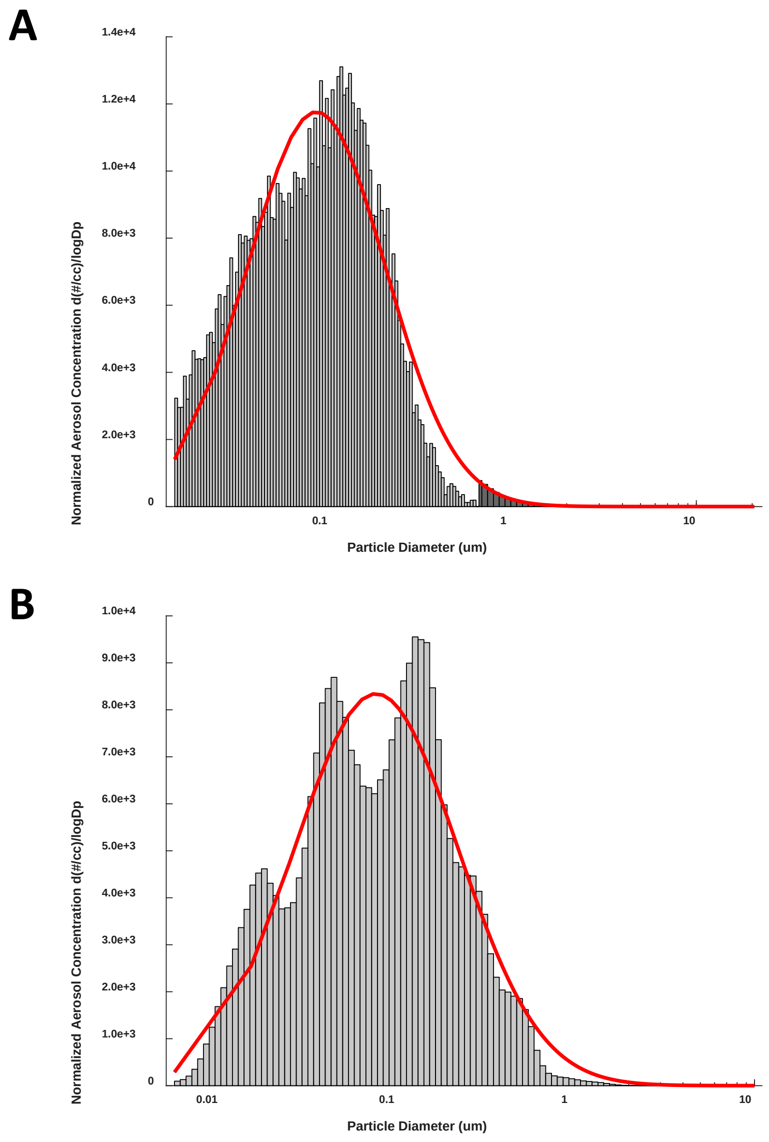


Figure S6: Saline particle size distributions sampled from the room during testing. A) Using a scanning mobility particle sizer (SMPS-light grey) and an aerodynamic particle sizer (APS-dark grey). The red line represents a log-normal fit of the histogram (count median diameter = 89.3 nm). B) Using a high resolution electrical low-pressure impactor (ELPI+) (count median diameter = 78.8 nm).

Particle size distribution measurements showed that particles spanned a wide range of sizes that cover much of the range of typical virus-laden aerosols (Fennelly 2020; Gralton et al. 2011). Non-negligible particle counts were registered from the bottom detectable ranges of the size analyzers (few nanometers) up to around 2 µm. Since the ranges of the SMPS and APS provide information about 2 separate size ranges (SMPS-nm range, APS- µm range) their data were combined and are shown in A. The data for the ELPI+ which covers the full range of aerosols measured is shown in B.

Table S1: Cough aerosol mass (µg) collected on each stage of the cascade impactor using the respiratory aerosol source control measurement system. The mass was adjusted for the amount of aerosol that was loaded into the system as described in Lindsley et al. (2020). SD is standard deviation.

| *Face covering* | *Replicates* | *Particle aerodynamic diameter (µm)* | | | | | | | | | | | |
|---|---|---|---|---|---|---|---|---|---|---|---|---|---|
| | | *0-0.6* | | *0.6-1.1* | | *1.1-2.1* | | *2.1-3.3* | | *3.3-4.7* | | *4.7-7* | |
| | | *Mean* | *SD* | *Mean* | *SD* | *Mean* | *SD* | *Mean* | *SD* | *Mean* | *SD* | *Mean* | *SD* |
| No face covering (control) | 26 | 89.75 | 11.72 | 94.09 | 9.56 | 194.14 | 18.15 | 101.73 | 26.95 | 34.05 | 5.77 | 10.00 | 1.85 |
| 3M 1860 N95 respirator | 6 | 4.71 | 0.88 | 1.30 | 0.20 | 0.89 | 0.18 | 0.18 | 0.04 | 0.08 | 0.01 | 0.06 | 0.00 |
| BYD N95 respirator | 4 | 22.84 | 0.74 | 20.75 | 0.80 | 36.24 | 1.95 | 8.95 | 0.86 | 1.76 | 0.13 | 0.46 | 0.04 |
| 3M 1818 surgical mask | 4 | 18.11 | 4.90 | 13.94 | 7.05 | 26.55 | 15.87 | 8.12 | 3.63 | 2.19 | 0.59 | 0.63 | 0.10 |
| Excel Artisan procedure mask | 4 | 50.46 | 5.95 | 51.31 | 2.13 | 90.20 | 6.13 | 28.56 | 2.50 | 7.18 | 1.45 | 1.83 | 0.19 |
| Hanes cloth mask | 6 | 62.34 | 7.44 | 56.64 | 6.14 | 92.41 | 9.43 | 31.39 | 3.40 | 8.63 | 1.03 | 2.29 | 0.39 |
| Enerplex mask | 4 | 70.61 | 3.93 | 59.20 | 4.45 | 101.77 | 6.87 | 32.37 | 5.18 | 7.31 | 0.96 | 2.01 | 0.23 |
| Craft & Soul mask | 4 | 63.58 | 2.89 | 60.62 | 2.02 | 101.28 | 6.97 | 34.96 | 1.93 | 11.14 | 1.45 | 3.79 | 0.64 |
| Lefty mask | 4 | 76.92 | 7.07 | 67.86 | 6.75 | 113.94 | 9.50 | 33.59 | 4.63 | 8.53 | 1.29 | 2.47 | 0.52 |
| Badger dust mask | 4 | 53.71 | 5.80 | 34.95 | 3.00 | 49.36 | 7.84 | 10.83 | 1.46 | 2.60 | 0.75 | 0.65 | 0.19 |
| Debrief Me mask | 4 | 67.14 | 7.83 | 66.39 | 9.41 | 122.45 | 29.61 | 35.49 | 7.98 | 6.83 | 0.92 | 1.57 | 0.17 |
| Fabrique Innovations mask | 4 | 53.25 | 5.29 | 47.91 | 5.92 | 67.81 | 9.18 | 17.51 | 3.87 | 5.43 | 0.82 | 1.64 | 0.24 |
| Besungo sports mask | 4 | 44.84 | 12.64 | 41.37 | 4.00 | 69.05 | 6.56 | 19.75 | 4.44 | 4.21 | 0.28 | 1.03 | 0.13 |
| Inspire mask w filter | 4 | 50.62 | 5.44 | 46.05 | 1.71 | 61.41 | 3.95 | 13.45 | 3.22 | 3.33 | 1.37 | 1.15 | 0.42 |
| FKGIONG gaiter 1-layer | 6 | 64.84 | 2.60 | 56.80 | 4.36 | 94.74 | 7.05 | 38.94 | 4.13 | 12.38 | 1.24 | 4.67 | 0.72 |
| FKGIONG gaiter 2-layer | 6 | 66.47 | 7.27 | 46.32 | 8.07 | 64.52 | 9.64 | 20.55 | 3.85 | 7.05 | 0.77 | 2.28 | 0.29 |
| AXBXCX gaiter 1-layer | 4 | 68.46 | 6.36 | 68.59 | 5.46 | 126.93 | 12.36 | 52.93 | 6.41 | 16.04 | 1.76 | 5.77 | 1.08 |
| AXBXCX gaiter 2-layer | 4 | 60.32 | 1.76 | 57.29 | 2.31 | 93.30 | 5.96 | 40.40 | 4.50 | 13.70 | 0.82 | 4.92 | 0.95 |
| Retro gaiter 1-layer | 4 | 61.79 | 3.82 | 57.92 | 13.87 | 96.53 | 20.81 | 37.00 | 5.32 | 11.67 | 1.98 | 4.02 | 0.91 |
| Retro gaiter 2-layer | 4 | 58.76 | 1.70 | 41.02 | 2.53 | 58.64 | 3.20 | 17.34 | 0.93 | 6.60 | 1.37 | 2.05 | 0.33 |
| Givon fleece gaiter | 4 | 57.09 | 3.22 | 48.46 | 3.05 | 84.73 | 8.28 | 36.27 | 4.34 | 12.52 | 1.44 | 4.69 | 0.60 |
| L&M bandana | 4 | 93.77 | 26.99 | 85.13 | 22.66 | 155.75 | 46.21 | 70.80 | 21.35 | 19.23 | 8.86 | 8.07 | 2.11 |
| Underwear bandana/gaiter | 4 | 62.68 | 12.92 | 68.25 | 4.76 | 118.63 | 2.68 | 42.57 | 1.22 | 13.71 | 0.60 | 4.70 | 0.72 |

Table S2: Breathing aerosol mass (μg) collected on each stage of the cascade impactor using the respiratory aerosol source control measurement system. The mass was adjusted for the amount of aerosol that was loaded into the system as described in Lindsley et al. (2020). SD is standard deviation.

| ***Face covering*** | ***Replicates*** | ***Particle aerodynamic diameter (μm)*** | | | | | | | | | | | |
|---|---|---|---|---|---|---|---|---|---|---|---|---|---|
| | | ***0-0.6*** | | ***0.6-1.1*** | | ***1.1-2.1*** | | ***2.1-3.3*** | | ***3.3-4.7*** | | ***4.7-7*** | |
| | | ***Mean*** | ***SD*** | ***Mean*** | ***SD*** | ***Mean*** | ***SD*** | ***Mean*** | ***SD*** | ***Mean*** | ***SD*** | ***Mean*** | ***SD*** |
| No face covering (control) | 35 | 79.55 | 8.66 | 88.45 | 7.57 | 188.49 | 19.28 | 93.91 | 9.55 | 35.27 | 3.77 | 11.15 | 1.39 |
| 3M 1860 N95 respirator | 4 | 1.00 | 0.40 | 1.04 | 0.72 | 2.05 | 1.57 | 0.75 | 0.54 | 0.24 | 0.18 | 0.09 | 0.04 |
| BYD N95 respirator | 4 | 4.95 | 0.68 | 5.03 | 0.52 | 9.45 | 1.05 | 3.42 | 0.24 | 0.87 | 0.09 | 0.28 | 0.03 |
| 3M 1818 surgical mask | 4 | 6.95 | 2.33 | 8.24 | 2.49 | 17.61 | 4.86 | 7.21 | 2.17 | 2.06 | 0.66 | 0.59 | 0.22 |
| Excel Artisan procedure mask | 4 | 47.59 | 2.23 | 55.38 | 5.12 | 115.30 | 11.29 | 52.68 | 5.04 | 13.84 | 1.23 | 4.00 | 0.39 |
| Hanes cloth mask | 4 | 51.28 | 9.47 | 61.42 | 14.83 | 111.54 | 29.02 | 40.89 | 9.22 | 9.84 | 2.98 | 2.58 | 0.54 |
| Enerplex mask | 4 | 38.72 | 2.50 | 48.08 | 3.90 | 94.03 | 12.33 | 38.23 | 7.16 | 13.33 | 3.83 | 3.73 | 1.36 |
| Craft & Soul mask | 4 | 49.60 | 14.01 | 50.82 | 10.63 | 96.11 | 22.11 | 34.90 | 9.15 | 9.09 | 1.60 | 2.52 | 0.44 |
| Lefty mask | 4 | 54.81 | 4.77 | 63.33 | 4.09 | 125.28 | 14.94 | 54.29 | 12.68 | 15.33 | 3.77 | 4.34 | 1.18 |
| Badger dust mask | 4 | 30.80 | 5.04 | 42.50 | 4.82 | 77.33 | 7.47 | 30.00 | 3.14 | 7.66 | 1.26 | 1.95 | 0.35 |
| Debrief Me mask | 4 | 35.74 | 15.19 | 39.27 | 13.61 | 71.44 | 26.76 | 27.37 | 14.79 | 6.85 | 3.83 | 1.69 | 0.89 |
| Fabrique Innovations mask | 4 | 35.42 | 5.27 | 38.02 | 2.15 | 70.21 | 7.18 | 20.43 | 3.30 | 4.98 | 1.40 | 1.21 | 0.36 |
| Besungo sports mask | 4 | 37.51 | 12.67 | 37.11 | 9.44 | 83.22 | 21.76 | 32.06 | 10.60 | 8.04 | 2.25 | 2.00 | 0.32 |
| Inspire mask w filter | 4 | 36.43 | 3.44 | 40.08 | 7.59 | 73.70 | 12.04 | 26.81 | 4.20 | 5.96 | 1.48 | 1.23 | 0.27 |
| FKGIONG gaiter 1-layer | 4 | 49.63 | 6.36 | 59.76 | 1.91 | 117.49 | 5.53 | 45.02 | 1.29 | 9.62 | 0.83 | 2.85 | 0.28 |
| FKGIONG gaiter 2-layer | 4 | 45.41 | 7.66 | 54.46 | 9.24 | 98.66 | 14.28 | 34.10 | 6.50 | 6.28 | 0.54 | 1.51 | 0.14 |
| AXBXCX gaiter 1-layer | 4 | 56.09 | 7.64 | 68.58 | 10.65 | 125.22 | 16.99 | 52.48 | 6.55 | 15.10 | 2.82 | 4.64 | 0.93 |
| AXBXCX gaiter 2-layer | 4 | 55.92 | 5.56 | 59.51 | 2.54 | 108.22 | 10.51 | 42.76 | 3.15 | 9.94 | 1.65 | 2.93 | 0.63 |
| Retro gaiter 1-layer | 4 | 42.70 | 9.36 | 46.01 | 6.80 | 87.66 | 9.46 | 33.88 | 2.04 | 7.54 | 0.53 | 2.17 | 0.16 |
| Retro gaiter 2-layer | 4 | 36.15 | 5.73 | 44.87 | 5.00 | 83.31 | 4.67 | 27.99 | 5.25 | 5.84 | 1.18 | 1.33 | 0.39 |
| Givon fleece gaiter | 4 | 61.00 | 10.54 | 59.58 | 10.32 | 94.01 | 9.37 | 22.38 | 0.93 | 4.24 | 0.71 | 1.06 | 0.09 |
| L&M bandana | 4 | 43.95 | 4.27 | 51.51 | 6.09 | 106.90 | 14.32 | 47.47 | 7.14 | 12.39 | 1.68 | 3.92 | 0.43 |
| Underwear bandana/gaiter | 4 | 53.48 | 8.36 | 62.95 | 11.12 | 127.18 | 19.21 | 51.40 | 5.68 | 13.65 | 1.31 | 3.71 | 0.26 |
